# Store-Operated Calcium Entry Controls Innate and Adaptive Immune Cell Function in Inflammatory Bowel Disease

**DOI:** 10.1101/2021.09.11.21263401

**Authors:** Marilena Letizia, Ulrike Kaufmann, Yin-Hu Wang, Lorenz Gerbeth, Annegret Sand, Max Brunkhorst, Jörn Felix Ziegler, Chotima Böttcher, Stephan Schlickeiser, Camila Fernández-Zapata, Kenneth Stauderman, Désirée Kunkel, IBDome researchers, Britta Siegmund, Stefan Feske, Carl Weidinger

**Author notes:** equal contribution (Marilena Letizia, Ulrike Kaufmann, Yin-Hu Wang, Stefan Feske, Carl Weidinger). Correspondence:* Stefan Feske, Department of Pathology, New York University Grossman School of Medicine, 550 First Avenue, New York, NY 10016, USA., Carl Weidinger, Charité - Universitätsmedizin Berlin, Campus Benjamin Franklin, Medizinische Klinik für Gastroenterologie, Infektiologie und Rheumatologie Hindenburgdamm 30, 12200 Berlin, Germany.

## Abstract

**Objective:** Inflammatory bowel disease (IBD) is characterized by dysregulated intestinal immune responses and constitutes a major clinical challenge in need of new treatment modalities to improve patient care. Store-operated Ca^2+^ entry (SOCE) is the predominant Ca^2+^ influx pathway in T cells and other immune cells, regulating many of their functional properties. It is currently unknown whether the pharmacologic blockade of SOCE represents a suitable drug-target for IBD treatment.

**Design:** Using mass and flow cytometry the effects of SOCE inhibition on lamina propria (LP) immune cells of patients with ulcerative colitis (UC) and Crohn’s disease (CD) were investigated. Primary organoid cultures served to study the impact of SOCE inhibition on the function, differentiation and survival of intestinal epithelial cells (IEC). T cell transfer models of colitis were applied to examine how the genetic or pharmacologic ablation of SOCE affects the clinical course of IBD in mice.

**Results:** We observed that the LP of IBD patients is characterized by an enrichment of innate lymphoid cells (ILC), CD4^+^ and CD8^+^ effector- as well as T regulatory cells producing IL-17 and TNFα. The pharmacologic inhibition of SOCE attenuated the production of pathogenic cytokines including IL-2, IL-4, IL-6, IL-17, TNFα and IFNγ by human colonic T cells and ILC, reduced the production of IL-6 by B cells and the production of IFNγ by myeloid cells, without affecting the viability, differentiation and function of primary IEC. T cell-specific genetic deletion of the SOCE signaling components *Orai1*, *Stim1* or *Stim2* revealed that the magnitude of SOCE correlates with the function of T cells and intestinal inflammation in mice. Moreover, the pharmacologic inhibition of SOCE alleviated the clinical course of colitic mice.

**Conclusion:** Our data suggest that SOCE inhibition may serve as a new pharmacologic strategy for treating IBD.

## Introduction

Inflammatory bowel disease (IBD) is a chronic inflammatory disease of the gastrointestinal (GI) tract that manifests predominantly as two related disease entities, ulcerative colitis (UC) and Crohn’s disease (CD). Both forms of IBD are associated with diarrhea, abdominal pain, fatigue as well as the development of colorectal cancer in patients with longstanding colitis and the development of fistulae and stricturing disease in patients with CD (1). Whereas UC predominantly affects the colon and is characterized by a superficial inflammation of the LP, CD can involve the entire GI tract and cause trans-mural inflammation. New single cell technologies such as single cell RNA-sequencing and mass cytometry have greatly advanced our pathophysiologic understanding of IBD (2, 3) as they have helped to better delineate the intestinal immune cell composition of IBD patients that drive intestinal inflammation. The LP of UC patients is characterized by a significant enrichment of TNFα producing CD8^+^ effector T cells (2), IL-17 producing effector memory CD4^+^ T cells and an expansion of T regulatory (Treg) cells producing inflammatory cytokines (3). HLA-DR^+^CD56^+^ granulocytes as well as TNFα− and INFγ−producing B cells are increased in the mucosa of CD patients (3). Despite these findings, the immune cell composition, signaling cascades and the cytokine networks controlling inflammation in therapy-refractory IBD remain incompletely understood. Recent advances that have revolutionized IBD treatment include blocking antibodies against pro-inflammatory cytokines such as TNFα (e.g. infliximab or adalimumab), IL-12 and IL-23 (ustekinumab) and against integrins such as vedolizumab (4). Nevertheless, intestinal and colonic resections are still frequently required in patients with therapy-refractory IBD, and novel treatment modalities are urgently needed to improve IBD outcomes.

SOCE is the predominant Ca^2+^ influx pathway in most immune cells and is required for the activation, differentiation and function of murine and human lymphocytes including B, NK and T cells. Stimulation of the T cell receptor (TCR) leads to production of the second messenger inositol-1,4,5 triphosphate (IP_3_), which triggers a transient release of Ca^2+^ from endoplasmic reticulum (ER) Ca^2+^ stores through IP_3_ receptors into the cytoplasm (5). The concomitant reduction of Ca^2+^ concentrations in the ER is sensed by stromal interaction molecules (STIM) 1 and STIM2 in the ER membrane, which are subsequently activated and translocate to the plasma membrane (6). Activated STIM1 and STIM2 bind to ORAI1 and its homologues ORAI2 and ORAI3, which form the pore of the Ca^2+^ release-activated Ca^2+^ (CRAC) channel. SOCE through CRAC channels causes a sustained elevation of intracellular Ca^2+^ levels (7, 8), which is required for the activation of numerous Ca^2+^-dependent enzymes including calcineurin, calmodulin kinases or Erk1/2 and transcription factors such as NF-κΒ, CREB and NFAT (9–11). SOCE is essential for the transcription of cytokines including IL-2, IFNγ and TNFα by T cells and other immune cells. In addition, SOCE regulates several metabolic pathways such as glycolysis and mitochondrial respiration, thereby controlling lymphocyte proliferation and effector functions (11–13). The pathophysiological importance of SOCE for immune function is emphasized by patients with loss-of-function mutations in *STIM1* or *ORAI1* genes, who suffer from combined immunodeficiency with recurrent infections (14–18). On the other hand, genetic or pharmacological inhibition of SOCE suppresses pro-inflammatory T cell functions and autoimmunity in animal models of multiple sclerosis and colitis (19–22).

Given the critical role of SOCE in immune cells, we hypothesized that pharmacologic blockade of SOCE might attenuate the pro-inflammatory function of lymphoid and myeloid immune cells from patients with therapy-refractory IBD and therefore represent a new strategy for immune modulation. Using T cell transfer models of colitis, we show that deletion of the CRAC channel genes *Stim1*, *Stim2* or *Orai1* in T cells prevents IBD and that the severity of intestinal inflammation correlates with the level of SOCE. By applying mass cytometry (CyTOF), we characterized immune cell subsets in the colonic LP of therapy refractory IBD patients and investigated the effects of the SOCE inhibitor BTP2 on intestinal immune cell populations from IBD patients. SOCE inhibition efficiently suppressed various pro-inflammatory functions of human T cells in a dosage dependent manner, inhibited the function of innate lymphoid cells (ILC) and, to a lesser degree, the function of myeloid cells and B cells. Importantly, treatment with the BTP2 had no detectable effects on the epithelial barrier functions of primary human and murine intestinal epithelial cells (IEC) *in vitro*. Treatment of mice in which IBD had been induced by adoptive T cell transfer with the selective SOCE inhibitor CM4620 attenuated the clinical course of colitis, which was associated with reduced neutrophil infiltration of the LP and decreased IFNγ and TNFα production by CD4^+^ T cells, suggesting that SOCE inhibition may be a new treatment option for IBD.

## Results

### SOCE in T cells is required for the induction of colitis in mice

We had previously shown that murine T cells lacking functional ORAI1 channels have reduced SOCE, resulting in impaired production of pro-inflammatory cytokines such as IFNγ and IL-17A, which was associated with a reduced capability to induce colitis upon their adoptive transfer into lymphopenic mice (20, 22). To better understand the role of other CRAC channel components in colitogenic T cells and the quantitative requirements of SOCE in T cell-mediated intestinal inflammation, we here used mice with T cell specific deletion of *Orai1*, *Stim1*, *Stim2* and both *Stim1/Stim2* genes. Naïve CD4^+^ T cells from *Orai1^fl/fl^Cd4*Cre, *Stim1^fl/fl^ Cd4Cre, Stim2^fl/fl^ Cd4Cre* and *Stim1^fl/fl^Stim2^fl/fl^Cd4Cre* mice were polarized into Th1, Th17 and induced Treg (iTreg) cells *in vitro* and analyzed for SOCE. Deletion of *Stim2*, *Orai1*, *Stim1* and both *Stim1/Stim2* resulted in a progressively more pronounced loss of SOCE (**Supplementary Figure 1A**). Deletion of CRAC channel components affected SOCE in Th1, Th17 and iTreg cells to a similar degree. Because of these graded effects on SOCE, we next isolated naive CD4^+^ T cells from these mice and injected them into *Rag1*^−/−^ host mice to induce colitis and to determine if the magnitude of SOCE determines the severity of intestinal inflammation (**Figure 1A**). *Stim2*-deficient CD4^+^ T cells induced weight loss and colitis (measured by histological scores) comparable to wild-type T cells (**Figure 1B,C**). By comparison, transfer of *Orai1*-deficient CD4^+^ T cells was associated with significantly attenuated weight-loss and disease severity. Transfer of *Stim1-*deficient T cells did not cause weight loss and no obvious signs of intestinal inflammation were detected (**Figure 1B,C**). Flow cytometric analysis of CD4^+^ T cells obtained from mesenteric lymph nodes (mLN) showed similar frequencies of cells producing IFNγ, TNFα and IL-17 when we compared *Stim2*-deficient and wild-type T cells (**Figure 1D**). By contrast, deletion of *Orai1* and *Stim1* resulted in significantly reduced frequencies of IFNγ, TNFα and IL-17 producing CD4^+^ T cells compared to wild-type cells. The frequencies of Treg cells were significantly reduced in the absence of *Orai1 -or Stim1,* but not in the absence of *Stim2* (**Figure 1D**). These findings suggested that moderate inhibition of SOCE in *Stim2-*deficient CD4^+^ T cells has no effects on T cell function, whereas more pronounced inhibition in *Orai1- or Stim1-*deficient CD4^+^ T cells gradually suppresses inflammatory cytokine production and the differentiation of iTreg cells *in vivo*. To determine the magnitude of SOCE required for T cell function and iTreg development, we differentiated *Orai1*, *Stim1*, *Stim2* and *Stim1/Stim2*-deficient naive CD4^+^ T cells into Th1, Th17 and iTreg cells *in vitro*. Deletion of *Stim2* had moderate effects on IFNγ and IL-17 production, whereas *Orai1*, *Stim1* and *Stim1/Stim2*-deficient Th1 and Th17 cells lacked production of these cytokines (**Figure 1E, Supplementary Figure 1B-C**). The numbers of Foxp3^+^ iTreg cells were not significantly reduced in the absence of *Stim2* expression, but were gradually more decreased by deletion of *Orai1*, *Stim1* and especially *Stim1/Stim2 (***Figure 1E**, **Supplementary Figure 1B-C**). When we correlated SOCE with cytokine production and Foxp3 expression, we found that inhibition of SOCE in murine T cells most profoundly suppresses IL-17 production followed by IFNγ production and to a lesser degree expression of Foxp3 (**Figure 1F, Supplementary Figure 1C**). These findings suggest that the amplitude of SOCE determines the fate of murine CD4^+^ T cells with the strongest dependence on SOCE observed in inflammatory Th17 and Th1 cells.

**Figure 1.**
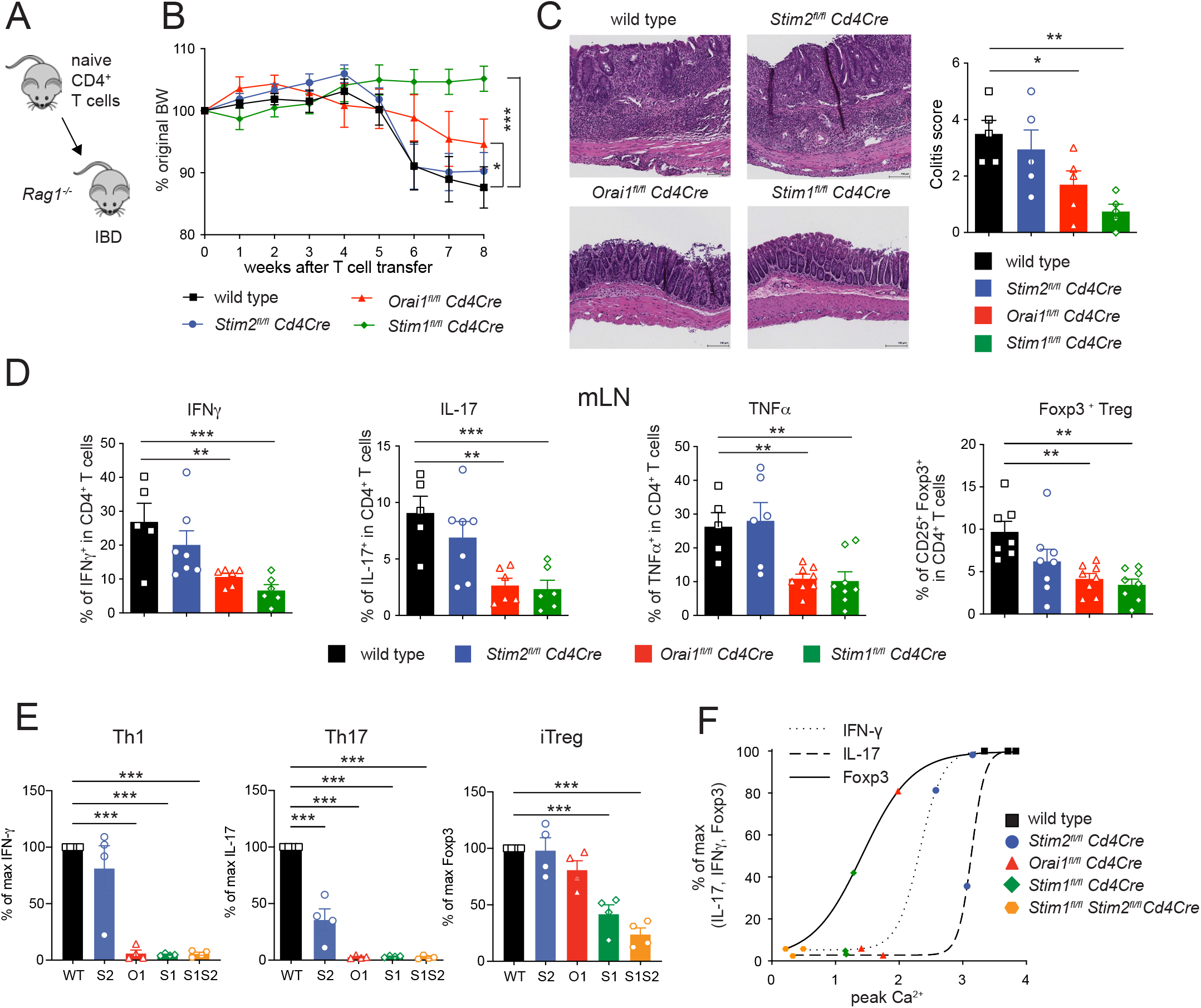
The genetic abrogation of Store-operated Calcium Entry (SOCE) prevents the development of intestinal inflammation in animal models of IBD. **(A-B)** Transfer model of colitis and weight curves of recipient *Rag1^−/−^* mice injected with either naive CD4^+^ T cells obtained from wild-type (wt), *Stim2^fl/fl^-Cd4-Cre*, *Orai1^fl/fl^-CD4-Cre* or *Stim1^fl/fl-^Cd4-Cre* mice. Summary of two independent experiments (n=3-5 animals per group) with a pooled total of 8 animals per group. (**C**) Exemplary H&E staining of colon tissue and histologic inflammation score 8 weeks after T cells transfer from 1 representative experiment, n=5 per group. (**D**) Cytokine production and frequencies of regulatory T cells in transferred T cells obtained from the mesenteric lymph node (mLN) of recipient *Rag1^−/−^* mice, data is pooled from 2 independent experiments with n=3-8 mice per group. (E) Signature protein expression for Th1 (IFNγ), Th17 (IL-17) and iTreg (Foxp3) lacking the SOCE signaling component *Stim1*, *Stim2*, *Orai1* or *Stim1* and *Stim2* subsets in relation to wild-type cells. Naïve CD4^+^ T cells were isolated from spleen and LNs of WT, *Stim2^fl/fl-^Cd4-Cre* (*S2*), *Orai1^fl/fl^-Cd4-Cre* (*O1*), *Stim1^fl/fl^-Cd4-Cre* (*S1*) and *Stim1^fl/fl^Stim2^fl/fl^-Cd4-Cre* (*S1S2*) deficient mice and differentiated into Th1, Th17 and iTreg cells, summary of 2 independent experiments, n=4 independent biologic replicates per condition and group. (**F**) Correlation of peak Ca^2+^ influx and relative expression of signature proteins in CD4^+^ T cell subsets. Statistical analyses by unpaired student’s t-test: ***p<0.001 **p<0.01 *p<0.05.

### Inhibition of SOCE gradually inhibits the production of pro-inflammatory cytokines by LPMCs in IBD patients

To investigate the role of SOCE in the function of lamina propria mononuclear cells (LPMCs) and intestinal inflammation in IBD patients, we isolated LPMCs from surgical colon resectates of IBD patients and peripheral blood mononuclear cells (PBMCs) from healthy donors (HD)(**Supplementary Table 1** for clinical details) and assessed Ca^2+^ influx by flow cytometry after *ex vivo* incubation with increasing concentrations (15 - 1,000 nM) of the CRAC channel inhibitor BTP2 (**Figure 2A**)(20, 23, 24). Treatment with BTP2 caused a dose-dependent inhibition of SOCE in CD4^+^ and CD8^+^ T cells with almost complete inhibition of SOCE inhibition at 1 μM (**Figure 2B**). 1 μM BTP2 also significantly reduced Ca^2+^ influx in PBMCs of HD, specifically in CD4^+^ and CD8^+^ T cells, CD19^+^ B cells, CD14^+^ monocytes and, to a lesser degree, CD56^+^ NK cells (**Supplementary Figure 2**), demonstrating that BTP2 is a potent inhibitor of SOCE in human gut-resident lymphocytes and PBMCs. To exclude that the observed differences in Ca^2+^ influx were caused by BTP2-induced cell death of lymphocytes, we measured apoptosis of LPMCs of IBD patients after incubation with 1 μM BTP2. As shown in **Supplementary Figure 3** we did not observe increased cell death in BTP2 treated LPMCs compared to untreated control cells.

**Figure 2.**
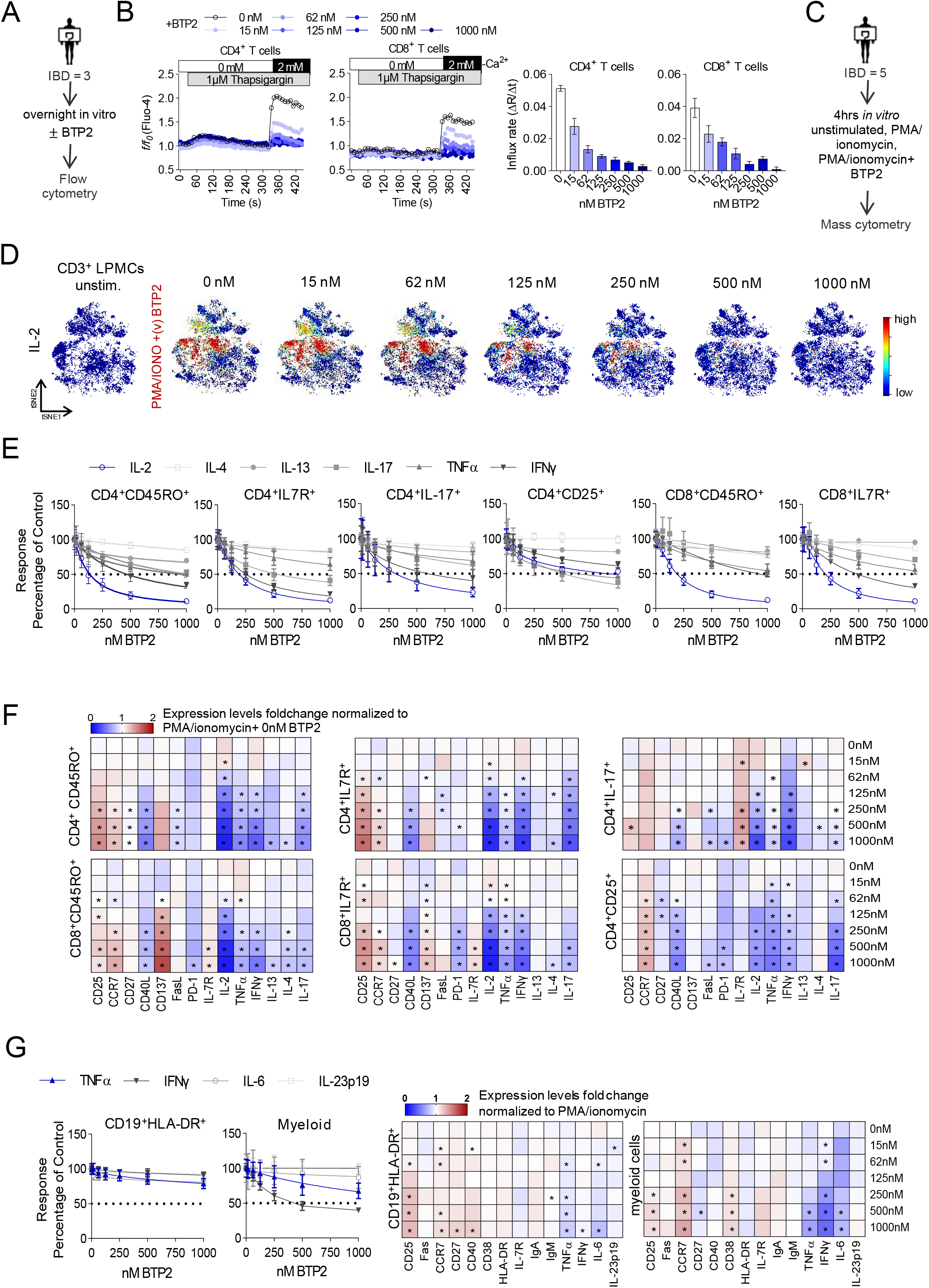
Gradual inhibition of Store-operated Calcium Entry (SOCE) reduces the production of pro-inflammatory cytokines in human lamina propria cells. (**A**) Experimental setup for Ca^2+^ influx assays on LPMCs (IBD: *n* = 3) by Flow cytometry. (**B**) Ca^2+^ influx rates in human lamina propria CD4^+^ and CD8^+^ T cells (IBD: *n* = 3) in the presence of 0-1000 nM BTP2 after stimulation with the sarco-endoplasmic reticulum Ca^2+^ ATPase (SERCA) inhibitor thapsigargin (TG). Bar plots showing the mean values. (**C**) Experimental setup for mass cytometry assay (CD: *n* = 5). (**D**) viSNE plots of CD45^+^CD3^+^ LPMCs (CD: *n* = 5) stimulated with PMA/ionomycin in the presence of 15-1000 nM BTP2 with un-stimulated samples serving as controls. viSNE plots are colored according to the expression level of IL-2 (blue: low, red: high) and are representative of one CD patient. (**E**) Dose response curve graphs reflecting the normalized production of cytokines in response to treatment with increasing dosages (0-1 µM) of BTP2 in CD45^+^CD3^+^ T cells activated 4 h *ex vivo* with PMA/ionomycin (CD: *n* = 5). The dose response was normalized to control samples treated with PMA/ionomycin. (**F**) Heatmaps representing the median fold change of cytokines and functional markers expression in CD45^+^CD3^+^ LPMCs (CD: *n* = 5) activated 4 h *ex vivo* with PMA/ionomycin ± 15 – 1000 nM BTP2, normalized to samples treated with PMA/ionomycin ± 15 – 1000 nM BTP2. Significant fold changes in marker expression levels between samples treated with vehicle (DMSO) alone and those treated with 15 - 1000nM BTP2 were calculated using a paired Wilcoxon matched-pairs signed rank test, *p <0.05. (**G**) Dose response curve graphs displaying the normalized cytokines production response to treatment with increasing dosages (15 – 1000 nM) of BTP2 in CD45^+^CD3^−^ T cells after 4h of ex vivo stimulation with PMA/ionomycin (CD: *n* = 5). The dose response was normalized to control samples stimulated with PMA/ionomycin. Error bars on all plots represent the standard mean error (SEM).

To determine the effects of SOCE inhibition on the function of LPMCs isolated from IBD patients, we subsequently evaluated the production of various pro-inflammatory cytokines and factors in LPMCs of 5 CD patients after BTP2 treatment (15 - 1,000 nM) *in vitro* by mass cytometry (**Figure 2C**). LPMCs were stained with a panel of antibodies against lineage markers of T cells (CD3, CD4, CD8), myeloid cells (CD11b, CD11c, CD14), B cells (CD19, IgM, IgA), and NK cells (CD56) similar to our previously published protocol (26). We furthermore included antibodies against differentiation and homing markers (CCR7, CD25, CD33, FAS (CD95), CD103, IL7R (CD127), CD137), activation markers (CD40, CD45RA, CD45RO, CD86, HLA-DR), cytokines (IL-2, IL-4, IL-6, IL-13, IL-17A, IL-23p40, IFNγ, TNFα, CD40L, FasL) and metabolic markers (CD27, CD38, PD-1). A detailed description of the antibody panel used for mass cytometry is provided in **Supplementary Table 2**.

To examine the effects of SOCE inhibition on T cell function in a BTP2 dose-dependent manner, we applied the t-distributed stochastic linear embedding (t-SNE) algorithm and the *FlowSOM/ConsensusClusterPlus* self-organizing map method (25), which allowed us to cluster CD45^+^CD3^+^ LPMCs into 9 major cell subtypes (**Supplementary Figure 4A-B**) according to the expression of major lineage markers listed in **Supplementary Table 3**. Subsequently, we compared the expression of cytokines and activation markers between LPMCs stimulated with PMA/ionomycin for 4h and samples treated with PMA/ionomycin in the presence of increasing concentrations of BTP2. Stimulation of LPMCs with PMA/ionomycin resulted in an increased of expression IL-2 in CD45^+^CD3^+^ LPMCs, which was inhibited gradually by increasing concentrations of the CRAC channel inhibitor BTP2 (**Figure 2D**). BTP2 also efficiently decreased the production of other cytokines including IFNγ, TNFα, IL-17, IL-13 and IL-4 in a dose-dependent manner (**Figure 2E**). Similar suppression of cytokine production was observed in effector and memory CD4^+^ and CD8^+^ T cells, and to a lesser extent in CD4^+^IL17^+^ Th17 cells. The weakest effects of SOCE inhibition on cytokine production were observed in CD4^+^CD25^+^IL7R^−^ Treg cells. Only IL-2, IL-17 and TNFα were suppressed by ∼ 50% with the highest concentration of BTP2 (1 μM) in Treg cells, whereas the same concentration of BTP2 strongly suppressed IL-2 and IFNγ in effector and memory CD4^+^ and CD8^+^ T cells. These findings are consistent with the greater resistance of murine iTreg cells to inhibition of SOCE (**Figure 1**)(5, 26) and further highlight the alternating requirements of cell-specific thresholds of SOCE signaling strength.

Besides cytokines, the expression of several functional markers of LPMCs such as CD40L, FasL and PD-1 was decreased by increasing concentrations of BTP2 (**Figure 2F**) which is consistent with the known role of Ca^2+^ and NFAT in their transcriptional regulation (27–29). For other markers such as CD25, CD137 (4-1BB) or CCR7, we found a dose-dependent up regulation upon treatment with BTP2 (**Figure 2F**). Interestingly, this increase was cell type specific and did not occur in all T cell subsets. For instance, CD25 was up regulated in BTP2-treated CD4^+^ and CD8^+^ effector and memory T cells, but not in Th17 and Treg cells after pharmacologic blockade of SOCE. Similarly, CD137 was only significantly up regulated after BTP2 treatment in effector and memory CD8^+^ T cells. These findings suggest that cell type-specific thresholds for the effects of SOCE may exist that control the functions of different T cell subsets.

To better understand the role of SOCE in other LPMC subsets besides T cells, we analyzed the expression of cytokines and functional markers in BTP2-treated CD45^+^CD3^−^ LPMCs of the same IBD patients (**Figure 2G, Supplementary Figure 5**). Surprisingly, cytokine production by CD19^+^ HLA-DR^+^ B cells was barely affected by increasing BTP2 concentrations. A slightly more pronounced suppression, especially of IFNγ and to a lesser degree of TNFα, was observed in CD11c^+^ myeloid cells. By contrast, SOCE inhibition resulted in a significant induction of CD25 and CCR7 expression in both B cells and myeloid cells (**Figure 2G**). It is noteworthy that the findings described above in LMPCs of CD patients could be confirmed in independent experiments using LPMCs isolated from 3 UC patients (**Supplementary Figure 6**). Collectively, these data demonstrate the significant effects of SOCE inhibition on pro-inflammatory cytokine expression in LP T cells, and to a lesser degree myeloid cells, of CD and UC patients and the complex, subset-dependent regulation of immune cell activation markers.

### Altered immune cell composition in the lamina propria of IBD patients

After establishing BTP2 as a potent inhibitor of SOCE and the function of human LPMC, we next investigated whether the function of disease-modifying LPMC subsets in IBD patients can be modulated by pharmacologic SOCE inhibition. To address this question, we first aimed to define UC- and CD-specific immune cell compositions in the LP of patients with therapy-refractory IBD, which is currently not well defined. We used the following stepwise approach: First, we compared the frequencies and functional status of immune cell subsets within LPMCs isolated from 6 patients with UC and 6 patients with CD undergoing colon resection due to refractory disease. Using mass cytometry, we compared the LPMC composition of IBD patients to that in non-inflamed colon mucosa obtained from non-IBD control patients (**Supplementary Table 1** for clinical details). Isolated LPMC were stimulated with PMA/ionomycin for 4h *ex vivo* and analyzed by CyTOF using a panel of 37 markers **(Figure 3A and Supplementary Table 2**), which allowed us to determine functional aspects of disease-specific cell subsets that drive inflammation in UC and CD. To investigate the effects of SOCE inhibition on these IBD-characterizing LPMC subsets, we stimulated LPMC in the presence or absence of a single concentration of 1µM BTP2 for which we had observed an almost complete suppression of Ca^2+^ influx and cytokine production in LP T cells (**Figure 2A**). For the evaluation of CyTOF data, we first performed a general high-dimensional analysis of all CD45^+^ cells that were clustered based on their expression of lineage markers (**Figure 3**). This was followed by in-depth analyses of CD45^+^CD3^+^ LP T cells (**Figure 4**) and CD45^+^CD3^−^ LPMCs (**Figure 5**) that were clustered according to their expression of surface makers and their expression of cytokines. For the above-mentioned analyses, we first compared the frequencies of cell clusters between UC, CD and non-inflamed controls and next assessed the effects of BTP2 on the activation and function of LPMC subsets characteristic of IBD.

**Figure 3.**
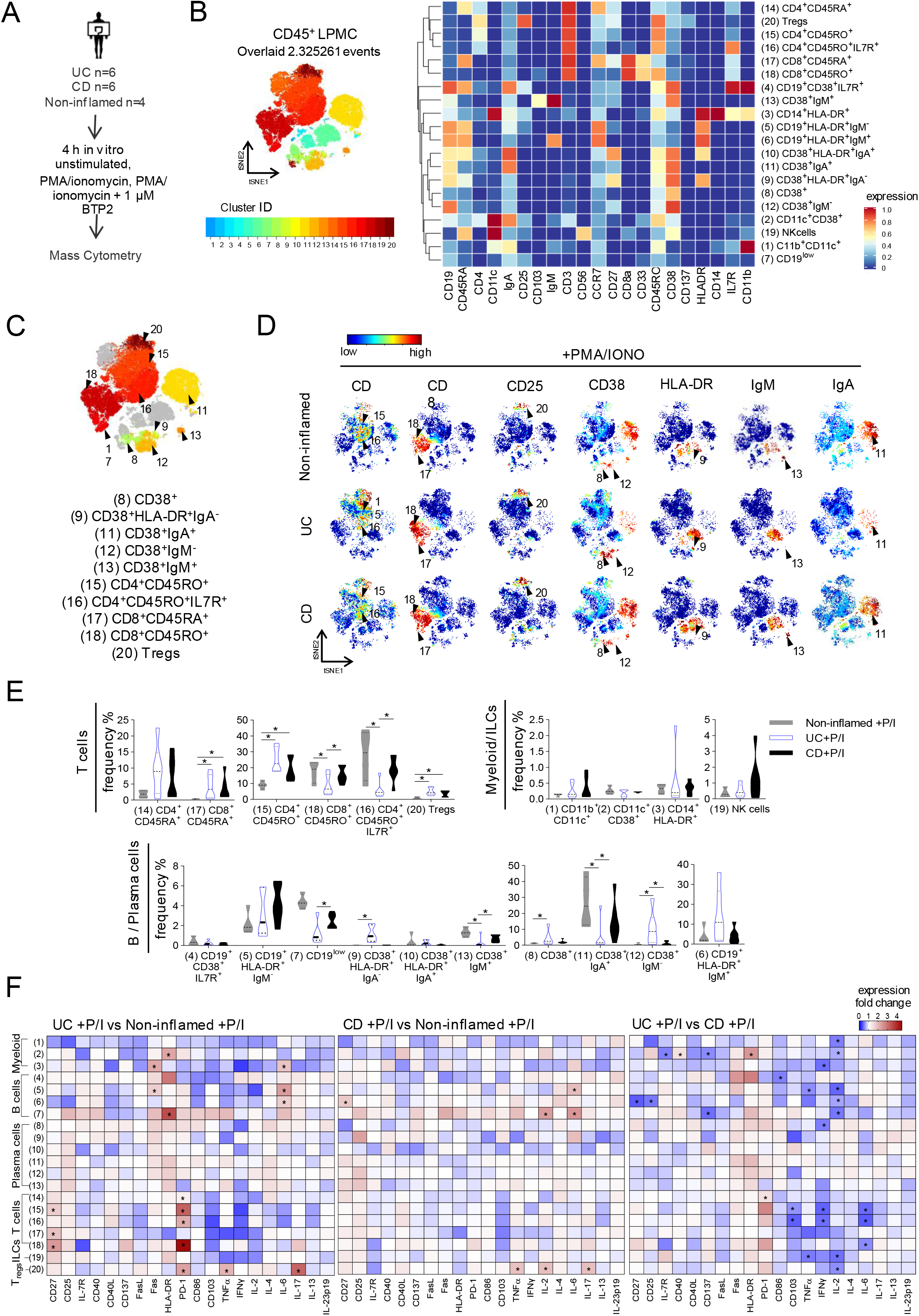
Global mass cytometric analysis of the immune cell composition in the colonic lamina propria obtained from therapy refractory IBD patients. (**A**) Experimental setup for mass cytometric assays (Non-inflamed: *n* = 4, CD: *n* =6, UC: *n* =6). (**B**) t-SNE plot of merged FCS files from LPMCs of Non-inflamed controls, UC or CD patients after ex-vivo stimulation with PMA/ionomycin ± 1µM BTP2. Colors on the t-SNE plot indicate 20 defined clusters representing major CD45^+^ LPMC-lineages. Heatmap clustering showing the expression levels of the 21 markers used for cluster analysis. (**C**) FlowSOM map of merged FCS files from CD45^+^ cells of Non-inflamed controls, UC and CD patients either left unstimulated or treated with PMA/ionomycin±1µM BTP2. The coloring indicates significant regulated cell clusters in CD patients vs. controls, UC patients vs. controls and between CD and UC patients. (**D**) viSNE plots of one exemplary Non-inflamed control, UC and CD patient colored by marker expression levels (blue: low, red: high). (**E**) Quantified frequencies (%) of each cell subset defined by the cluster analysis. The star (*) depicts significant differences between Non-inflamed controls vs UC, Non-inflamed and CD or UC and CD samples stimulated 4h *in vitro* with PMA/ionomycin. Statistics were calculated using the edgeR statistical framework with negative binomial GLM and a false discovery rate adjusted to 10%, BH. (**F**) Heatmaps representing the medianfold change of cytokines and functional markers expression in CD45^+^ LPMCs activated 4 h *ex vivo* with PMA/ionomycin among Non-inflamed, UC and CD samples. Significant differences were calculated by a Two-stage linear step-up procedure of Benjamini, Krieger and Yekutieli, with Q = 1% and are indicated by stars (*).

**Figure 4.**
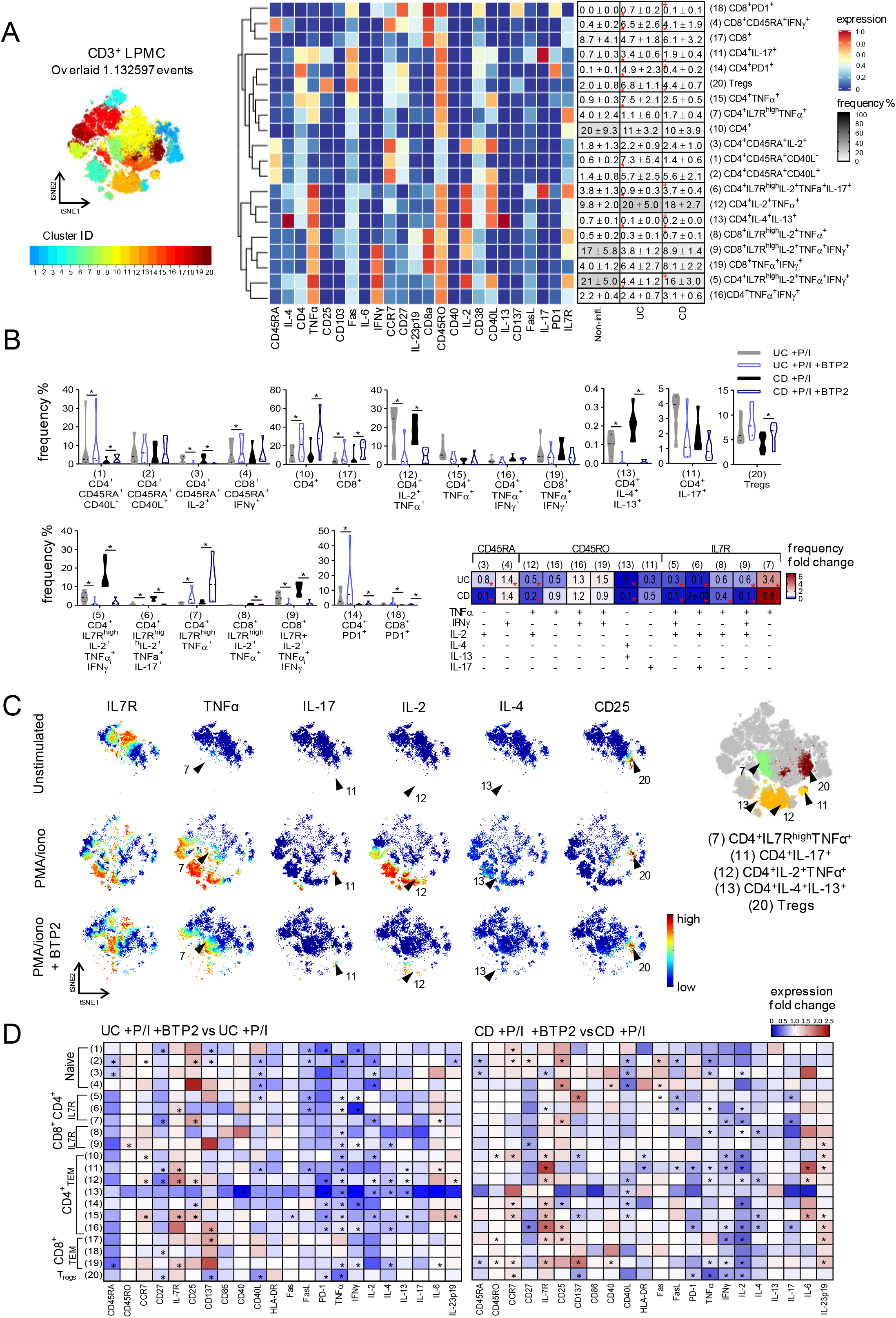
Inhibition of SOCE blocks the activation and function of human colonic T cells. (**A**) t-SNE plot of merged FCS files from samples treated with PMA/ionomycin ± 1µM BTP2 (Non-inflamed: *n* = 4, CD: *n* =6, UC: *n* =6). Colors on the t-SNE plot indicate 20 defined clusters among CD45^+^CD3^+^ LPMCs. Heatmap clusters show the expression levels of the 24 markers used for cluster analysis. The table on the right displays the mean values (±SEM) of quantified frequencies (%) for each cell subset defined in Non-Inflamed controls, UC and CD patients after 4h *in vitro* stimulation with PMA/ ionomycin. Significant differences were calculated using the edgeR statistical framework with negative binomial GLM and a false discovery rate adjusted to 10%. (*) indicates statistical significant differences between UC and Non-inflamed patients or CD and Non-inflamed patients, (^+^) indicates significant differences between UC and CD patients. **(B)** Violin plots showing quantified frequencies (%) of each cell subset defined by the cluster analysis in Non-Inflamed, UC and CD patients stimulated 4h *in vitro* with PMA/ionomycin±BTP2.Significant differences between UC+PMA/ionomycin and UC+PMA/ionomycin+BTP2 or CD+PMA/ionomycin and CD+PMA/ionomycin+BTP2 were calculated by a paired Wilcoxon matched-pairs signed rank test, *p < 0.05. The table on the right summarizes CD45^+^CD3^+^ clusters producing at least one cytokine in CD45RA naïve, CD45RO effector and IL7R memory cells. The table shows the median of frequency fold change in UC+PMA/ionomycin+BTP2 or CD+PMA/ionomycin+BTP2, normalized to UC+PMA/ionomycin or CD+PMA/ionomycin respectively. **(C)** viSNE plots of one exemplary CD patient colored by marker expression levels (blue; low, red: high). **(D)** Heatmaps indicating the median fold change of cytokines and functional markers expression in CD45^+^CD3^+^ LPMCs activated 4 h *in vitro* with PMA/ionomycin± BTP2, normalized to UC+PMA/ionomycin or CD+PMA/ionomycin respectively. Significant differences were calculated by a paired Wilcoxon matched-pairs signed rank test, *p <0.05.

**Figure 5.**
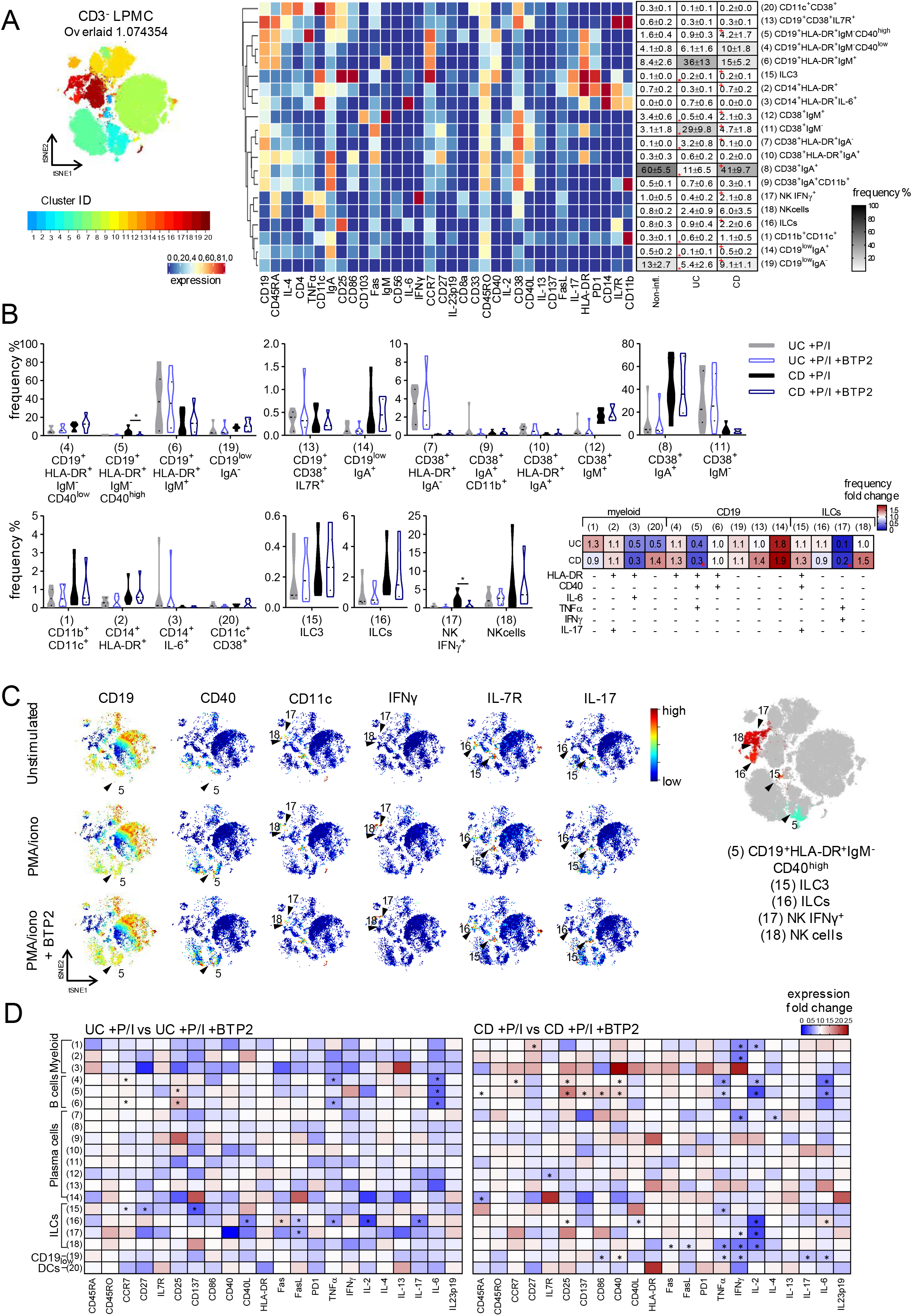
Effects of SOCE inhibition on CD3^−^ lamina propria cells. (**A**) t-SNE plot of merged FCS files from unstimulated samples or samples treated with PMA/ionomycin ± 1µM BTP2 (Non-inflamed: *n* = 4, CD: *n* =6, UC: *n* =6). Colors on the t-SNE plot indicate 20 defined clusters among CD45^+^CD3^−^ LPMCs. Heatmap clusters showing the expression levels of the 34 markers used for cluster analysis. The table on the right displays the mean values (±SEM) of quantified frequencies (%) for each cell subset defined in Non-Inflamed, UC and CD patients after 4 h stimulation with PMA/ionomycin. Significant differences were calculated using the edgeR statistical framework with negative binomial GLM and A false discovery rate adjusted to 10%, BH (*) indicates statistical significant differences between UC and Non-inflamed patients or CD and Non-inflamed patients, (^+^) indicates statistical significant differences between UC and CD patients. **(B)** Violin plots displaying quantified frequencies (%) of each cell subset defined by the cluster analysis in samples of Non-Inflamed controls, UC or CD patients after ex-vivo stimulation (4h) with PMA/ionomycin ± BTP2. Significant differences between UC+PMA/ionomycin and UC+PMA/ionomycin+BTP2 or CD+PMA/ionomycin and CD+PMA/ionomycin+BTP2 were calculated by a paired Wilcoxon matched-pairs signed rank test, *p < 0.05. The table on the right summarizes CD45^+^CD3^−^ clusters defined as myeloid, CD19^+^ B cells or ILCs. Table showing the median of frequency fold change in UC+PMA/ionomycin+BTP2 or CD+PMA/ionomycin+BTP2, normalized to UC+PMA/ionomycin or CD+PMA/ionomycin respectively. Significant differences were calculated by a paired Wilcoxon matched-pairs signed rank test, *p <0.05. **(C)** viSNE plots of one exemplary CD patient colored by marker expression levels (blue; low, red: high). **(D)** Heatmaps indicating the median fold change of cytokines and functional markers expression in CD45^+^CD3^−^ LPMCs activated 4 h in vitro with PMA/ionomycin±BTP2, normalized to UC+PMA/ionomycin or CD+PMA/ionomycin respectively. Significant differences were calculated by a paired Wilcoxon matched-pairs signed rank test, *p <0.05.

The analysis of all CD45^+^ cells (**Supplementary Figures 6**) allowed us to identify 20 clusters of LPMC subsets based on the expression of cell lineage markers listed in **Supplementary Table 4** and **Figure 3B**. LPMC clusters that are significantly dysregulated in UC and/or CD patients compared to non-inflamed controls are highlighted on the CD45^+^ FlowSOM map in **Figure 3C** and complementary viSNE maps for individual markers are shown in **Figure 3D**.

This analysis revealed several significant differences between LPMC populations in CD patients vs. controls, UC patients vs. controls and between CD and UC patients: Compared to LPMC from non-inflamed controls, UC and CD patients both showed a significant increase in the frequencies of CD4^+^CD45RO^+^ effector T cells (cluster 15), CD8^+^CD45RA^+^ naive T cells (cluster 17) and CD4^+^CD25^+^IL7R^−^ Treg cells (cluster 20) **(Figure 3E)**. Increased Treg frequencies are consistent with previous reports (30, 31) and are in line with the hypothesis that Treg numbers are elevated in the LP of IBD patients as a compensatory immune-regulatory mechanism to control the ongoing inflammation. We also observed differences specific to UC patients: Thus, the frequencies of CD38^+^IgA^+^ (cluster 11) and CD38^+^IgM^+^ (cluster 13) plasma B cells were significantly reduced in UC patients compared to CD patients and non-IBD controls, whereas their CD38^+^ (cluster 8), CD38^+^HLA-DR^+^IgA^−^ (cluster 9) and CD38^+^IgM^−^ (cluster 12) B cells were significantly expanded compared to non-inflamed controls, suggesting a profound, UC specific dysregulation of the B cell compartment. UC patients also had significantly fewer CD4^+^CD45RO^+^IL7R^+^ effector memory T cells (cluster 16) and CD8^+^CD45RO^+^ effector T cells (cluster 18) compared to CD patients and controls.

After defining the differences between LPMC subsets in IBD patients and controls, we next analyzed the expression levels of activation markers and cytokines in LPMC of UC, CD patients and non-inflamed controls within these cell clusters after stimulation of LPMC with PMA/ionomycin for 4h *ex vivo* (**Figure 3F**). Intriguingly, Treg cells (cluster 20) of both UC and CD patients produced more pro-inflammatory cytokines such as TNFα and IL-17 compared to non-inflamed controls, whereas only Treg cells of CD patients also showed increased production of IL-2. These findings are in line with a recent report by Mitsialis et al (3). Furthermore, we observed a significant up regulation of PD-1 and CD27 on different T cell subsets of UC but not CD patients compared to controls, suggesting that T cell exhaustion might be a feature of the chronic inflammatory milieu in UC patients. Myeloid cells and B cells of both UC and CD patients were characterized by a higher expression of HLA-DR and IL-6 compared to non-inflamed controls. Overall, these findings demonstrate that the balance of effector CD4^+^ and CD8^+^ T cells in the LP of IBD patients is severely perturbed compared to non-inflamed controls. Of note, our findings highlight a specific role of PD-1 expressing exhausted T cells in UC but not CD patients. In addition, we observed profound differences in the composition of humoral and innate immune cells in the LP of UC and CD patients, with a particular involvement of CD38^+^ plasma cells producing IgA and IgM in CD but not UC patients.

### Inhibition of SOCE modulates the function of IBD-defining colonic T cell populations

We next aimed to determine in more detail how SOCE regulates the activation of IBD-characterizing LPMCs and their ability to produce pro-inflammatory cytokines that are implicated in the pathophysiology of UC and CD (8). We therefore conducted separate analyses for CD45^+^CD3^+^ T cells (**Figure 4**) and CD45^+^ CD3^−^ non-T cells (**Figure 5**) within LPMC of IBD patients and non-inflamed controls using the gating strategy shown in **Supplementary Figure 7-8**. By subsequently clustering the cells not only based on their expression of lineage markers but also by their expression of differentiation markers and cytokines (**Supplementary Table 5)** we identified 20 subsets of cells within CD45^+^CD3^+^ LP T cells (**Figure 4A**). UC and CD patients were both characterized by an enrichment of CD4^+^IL-17^+^ (cluster 11), CD8^+^CD45RA^+^IFNγ^+^ T cells (cluster 4) and CD4^+^ Treg cells (cluster 20) compared to non-inflamed controls (**Figure 4A**). Unexpectedly, the frequency of CD4^+^IL-4^+^IL-13^+^ Th2 cells (cluster 13) was significantly reduced in both CD and UC compared to control samples, suggesting that these cells may not play a major pathogenic role in the maintenance of chronic inflammation in therapy-refractory IBD. UC patients harbored higher frequencies of CD4^+^ PD1^+^ (cluster 14), CD8^+^ PD1^+^ (cluster 18) and CD4^+^ TNFa^+^ (cluster 15) T cells when compared to CD patients and non-IBD controls. By contrast, the frequencies of CD4^+^ IL7R^high^ IL-2^+^ TNFα^+^ IFNγ^+^ (cluster 5) and CD4^+^ IL7R^high^ IL-2^+^ TNFα^+^ IL-17^+^ (cluster 6) T cells were significantly reduced in UC patients compared to CD patients and non-inflamed control samples (**Figure 4A**). Together these findings demonstrate that the composition of CD4^+^ and CD8^+^ T cell populations and their activation status is significantly perturbed in the LP of IBD patients with further differences between UC and CD patients.

We next investigated how the inhibition of SOCE affects the function of the identified CD3^+^ T cell subsets isolated from the LP of UC and CD patients. We paid particular attention to differential effects of SOCE inhibition on specific T cells subsets in order to discriminate the individual role of SOCE in each of the described T cell subsets. Inhibition of SOCE by 1 μM BTP2 during *ex vivo* stimulation with PMA*/*ionomycin resulted in strongly reduced abundances of naive CD4^+^IL-2^+^ T cells (cluster 3), CD4^+^IL-2^+^TNFa^+^ (cluster 12) and IL-4^+^IL-13^+^ Th2 cells (cluster 13) in UC and CD patients **(Figure 4B,C)**. Moreover, the frequencies of pro-inflammatory CD4^+^IL-7R^+^ and CD8^+^IL-7R^+^ memory T cells that produced IL-2, IFNγ and TNFα (clusters 5 and 9) were significantly reduced in UC and CD patients after SOCE inhibition. Likewise, we observed a decrease in the percentages of CD4^+^IL-17^+^ T cells (cluster 11) and CD4^+^IL-7R^+^ memory T cells producing IL-2, TNFα and IL-17 (cluster 6) in BTP2-treated LPMC of UC and CD patients.

Inhibition of SOCE in LPMC isolated from UC and CD patients resulted in parallel, in a relative increase of non-activated CD4^+^ and CD8^+^ T cells (clusters 10 and 17cells) and naive CD4^+^ T cells (cluster 1) as well as in an increase of cells that were characterized by low or intermediate TNFα expression such as CD4^+^IL-7R^high^ TNFα^+^ memory T cells (cluster 7) (**Supplementary Figure 9**, **Figure 4B,C**). We attribute this increase to the fact that BTP2 potently suppresses T cell activation and TNFα expression, thereby increasing the frequencies of T cells that are not activated and that are intermediate producers of TNFα. BTP2 treatment also increased the frequencies of CD4^+^PD1^+^ and CD8^+^PD1^+^ T cells (clusters 14 and 18) in both UC and CD patients (**Figure 4B, C**) and resulted in a relatively increased abundance of CD4^+^CD25^+^IL7R^−^ Treg cells (cluster 20) in CD patients.

We next determined the effects of SOCE inhibition on the expression levels of individual cytokines and functional markers of T cells **(Figure 4D)**. Treatment with BTP2 resulted in a significant suppression of TNFα in the majority of T cell subsets of UC patients (**Figure 4D**, left panel) and to a lesser degree in T cells of CD patients (**Figure 4D**, right panel). IFNγ and IL-2 production were similarly affected by BTP2 in UC and CD patient samples; a notable exception were Treg cells because IFNγ and IL-2 production were only significantly suppressed by BTP2 in patient samples of CD but not UC. The effects of SOCE inhibition on IL-4, IL-13 and IL-17 production were more moderate and reached significance in only a few T cell subsets in both UC and CD patients. It is noteworthy, however, that BTP2 inhibited the production of cytokines and suppressed the expression of activation markers in the IL-4^+^IL-13^+^ Th2 subset (cluster 13) in UC patients and to a lesser degree also in CD patients, suggesting that SOCE is a potent regulator of Th2 cell function. Among surface markers, we observed a significant decrease in the expression of FasL, CD40L and PD-1 upon inhibition of SOCE, which was expected since the transcription of these molecules, like that of many of the cytokines described above, is dependent on Ca^2+^ signaling and NFAT (27–29). In contrast, the increased expression of several T cell markers such as IL-7R, CD137, CD25 and CCR7 as well as the cytokine IL-23p19 in certain T cell subsets of UC and CD patients after BTP2 treatment was not anticipated. It is noteworthy that the expression of the co-stimulatory receptor CD137 (4-1BB) was differentially affected by SOCE inhibition; while it was up-regulated in BTP2-treated effector T cells, its expression decreased in Treg cells upon inhibition of SOCE. Collectively, our data demonstrate that SOCE is a potent regulator of T cell activation and function in LP T cells of both UC and CD patients as the pharmacologic blockade of SOCE effectively suppresses the production of pro-inflammatory cytokines, in CD4^+^ effector and memory T cells in the LP of UC and CD patients.

### SOCE regulates the function of NK cells and ILCs and is required for the activation of B cells

We next focused our analysis on characterizing the composition of non-T cell subsets within LPMC of IBD patients and non-inflamed controls and investigated the effects of SOCE inhibition on these cells. In-depth analysis of CD45^+^CD3^−^ cells allowed us to define 20 cell clusters (**Figure 5A)** based on the expression of lineage markers, cytokines and functional markers described in **Supplementary Tables 2 and 6**. We detected an increase of IL-17 producing ILC3 cells (cluster 15) in colonic mucosa samples of UC and CD patients compared to non-inflamed tissue controls (**Figure 5A**), which is in line with a recent study that had observed a larger abundance of ILC3 cells in the peripheral blood of IBD patients with active disease (3). In addition, the frequencies of CD11c^+^CD11b^+^ myeloid cells (cluster 1) were increased in UC samples and, to a lesser degree, in CD patients compared to non-inflamed controls. Interestingly, CD samples harbored increased frequencies of CD3^−^CD19^−^Il7R^+^CD25^+^ ILCs (cluster 16) and IFNγ^+^ NK cells (cluster 17) as well as activated HLA-DR^+^CD40^+^ B cells (cluster 5) compared to UC samples. A notable difference between both patient cohorts was the decrease in class switched CD38^+^IgA^+^ B cells (cluster 8) and increase in CD38^+^IgM^−^ B cells (cluster 11) in UC compared to CD patients and healthy controls (**Figure 5A**).

To test the effects of SOCE inhibition on the function of non-T cells in the LP of IBD patients we stimulated CD45^+^CD3^−^ LPMC isolated from UC and CD patients *ex vivo* with PMA/ionomycin in the presence or absence of BTP2 **(Figure 5B, C)**. SOCE inhibition significantly decreased the frequencies of IFNγ^+^CD56^+^ NK cells (cluster 17) and activated HLA-DR^+^CD40^+^ B cells (cluster 5) in CD, but not in samples obtained from UC patients. No notable effects of SOCE inhibition on other cell clusters, including B cells, myeloid cells and NK cells were observed.

In analogy to our previous analyses, we subsequently analyzed how BTP2 treatment affects the expression of pro-inflammatory cytokines by non-T cells in the LP. Several subsets of activated CD19^+^HLA-DR^+^ B cells (clusters 4, 5 and 6) displayed a strong reduction of IL-6 production in samples of UC and CD patients treated with BTP2 (**Figure 5D**). In UC patients, the production of TNFα, IL-2 and IL-17 and the expression of CD40L and FasL was significantly reduced in ILCs (cluster 16) after exposure to BTP2 (**Figure 5D**). UC patients furthermore featured decreased levels of CCR7, CD27 and CD137 expression in ILC3 cells (cluster 15). In CD patients, BTP2 suppressed the production of TNFα, IL-2 and IFNγ as well as the expression of FasL by NK cells (cluster 18). Inhibition of SOCE furthermore significantly reduced the expression of IFNγ by myeloid cells of CD patients (**Figure 5D**). By contrast, the activation markers CD25, CD137, CD86 and CD137 were significantly up-regulated on CD19^+^ B cells (cluster 5) after BTP2 treatment. Taken together, inhibition of SOCE suppressed the expression of several inflammatory cytokines including IL-6 (B cells), TNFα (ILC and NK cells) and IFNγ (myeloid and NK cells) in IBD patients, suggesting that SOCE contributes to the pro-inflammatory milieu in the LP of IBD patients generated by myeloid cells and ILCs. Compared to the pronounced effects of BTP2 on CD3^+^ T cells **(Figure 4)**, the inhibition of SOCE had more moderate effects on CD3^−^ LPMC, suggesting that Ca^2+^ signals are more consequential for intestinal T cells than non-T cells.

### SOCE inhibition does not affect the homeostasis of primary murine and human intestinal epithelial cells

After establishing that genetic deletion of CRAC channel components in mouse T cells **(Figure 1)** and pharmacological suppression of SOCE in human LPMC **(Figures 3-5)** attenuates the pro-inflammatory function of T cells, B cells, myeloid and innate immune cells, we investigated how SOCE inhibition affects intestinal epithelial cells (IEC). Surprisingly, very limited information is available about the physiological role of CRAC channels in IEC function (32). To investigate how SOCE inhibition affects the survival, differentiation and function of IEC, we first isolated colonic crypts from C57BL/6 wild-type mice, which were cultured *ex vivo* into colonic organoids or 2D organoids monolayers. Treatment of murine IEC-derived organoids with 1 µM BTP2 did not significantly impact their viability (**Figure 6A, B**). Moreover, the trans-epithelial resistance of primary murine colonic epithelial cells that were grown in in 2D layers did not change after treatment with 1 µM BTP2 compared to DMSO-treated controls (**Figure 6C**). To validate these results in human IEC, we isolated primary human epithelial crypts from surgical colon specimen of UC patients and cultured them into organoids and 2D monolayers. Treatment of human IEC organoids with 1 µM BTP2 did not affect their viability **(Figure 6D)**. In analogy to our findings in murine IEC, BTP2 did not alter the trans-epithelial resistance of human IEC grown in 2D layers **(Figure 6E)**. Likewise, the differentiation of human IEC was not influenced by BTP2 treatment as no significant differences in the mRNA expression of IEC differentiation markers ALPI, MUC2 and CHGA were observed compared to DMSO treated controls **(Figure 6E)**. Together, these data indicate that the same concentrations of 1µM BTP2 that are sufficient to inhibit the activation and pro-inflammatory functions of LPMCs do not interfere with the differentiation, viability and barrier function of IEC.

**Figure 6.**
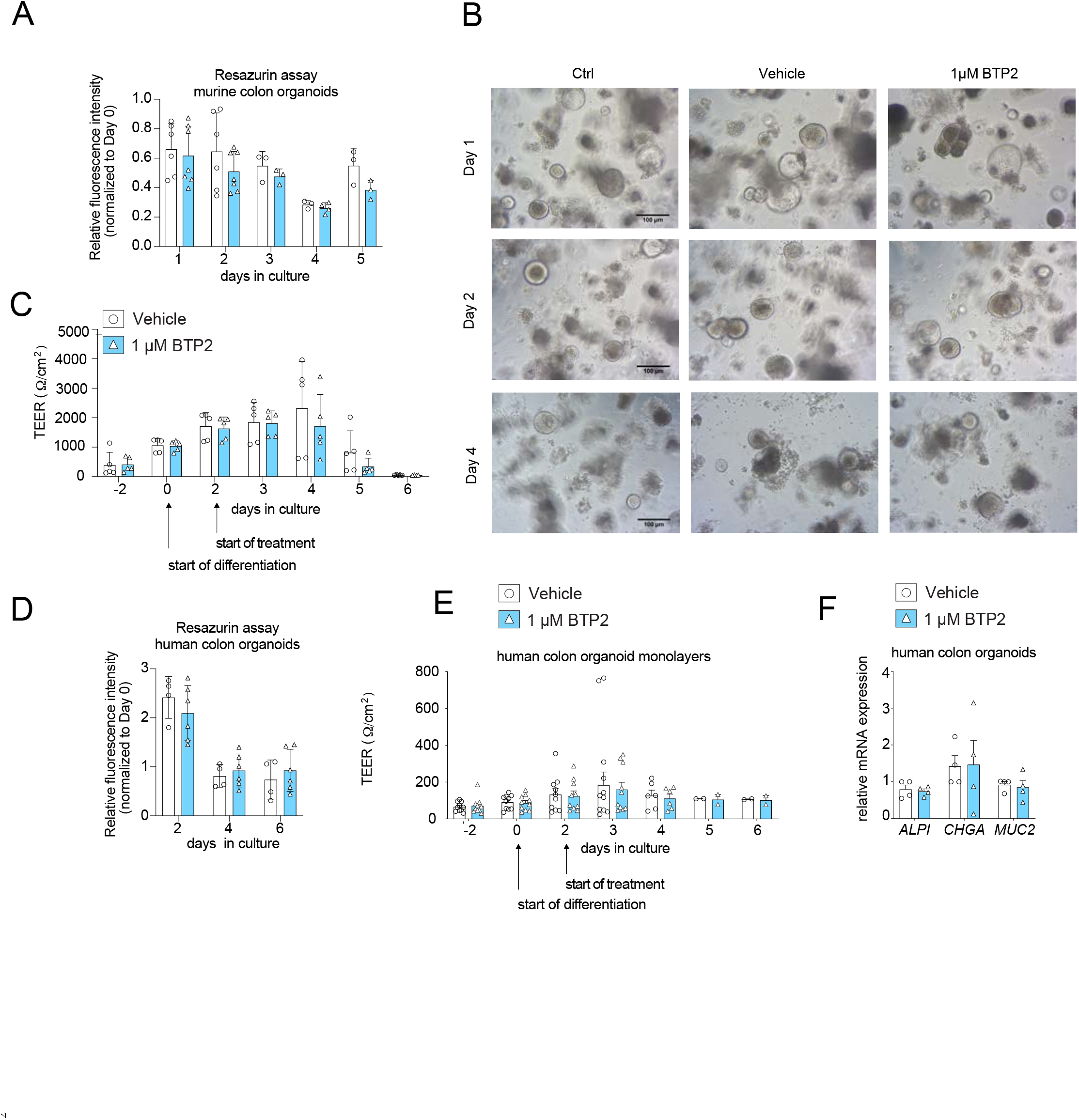
Treatment of murine and human intestinal epithelial cells with 1 µM BTP2 does not interfere with their survival, differentiation and function. (**A-B**) Adult stem cell-derived intestinal epithelial spheroids of wild-type C57Bl/6 mice were cultured for 4 days in Wnt-free medium to induce differentiation. Initial viability was determined on day 0. BTP2 or vehicle control (DMSO) was added to the culture medium and viability was measured after every 24h by resazurin viability assays and microscopy. (**C**) Transepithelial electrical resistance (TEER) was measured on differentiated murine colon monolayers in the presence or absence of 1µM BTP2. (**D**) Human colonic spheroids were generated from primary human epithelial crypts obtained from colon resectates of 2 UC patients and subsequently cultured for 6 days in Wnt-free medium to induce differentiation into colonic organoids. Initial viability was determined (Day 0). 1 µM BTP2 or vehicle control (DMSO) was added to the culture medium and viability was measured after 2, 4 or 6 days. (**E**) TEER was measured on differentiated human colon monolayers in the presence or absence of 1 µM BTP2. (**F)** Relative mRNA expression of markers for enterocytes (*ALPI*), goblet cells (*MUC2*), and entero-endocrine cells (*CHGA*) was quantified by RT-PCR in differentiated human colon monolayers after addition of 1 µM BTP2. All figures showing data of at least 2 independent experiments with 2-4 replicates for every condition tested.

### Pharmacological inhibition of SOCE *in vivo* ameliorates IBD and suppresses proinflammatory cytokine production by T cells

Because of the potent suppressive effects of SOCE inhibition with BTP2 on the expression of proinflammatory cytokines and activation markers by human LPMCs and the reduced ability of *Orai1* and *Stim1*-deficient T cells to induce IBD in mice, we investigated if the systemic application of a selective CRAC channel inhibitor is able to alleviate intestinal inflammation and the course of IBD in mice. In contrast to the adoptive transfer of *Orai1* and *Stim1*-deficient T cells into *Rag1^−/−^* mice, in which T cells lack SOCE from the start of the experiment, injection of mice with a CRAC channel inhibitor starting 18 days after T cell transfer tests whether SOCE inhibition is able to alter the progression of established disease rather than preventing it (**Figure 7A**). For these experiments we used the selective CRAC channel blocker CM4620, which can be formulated for *in vivo* use and is currently in phase 2 clinical trials for COVID-19 associated pulmonary inflammation and acute pancreatitis (NCT04195347). Transfer of naïve CD4^+^ T cells induced weight loss in recipient *Rag1^−/−^* mice that were gavaged with vehicle alone, whereas mice treated with CM4620 showed moderate gain of weight (**Figure 7B**). Systemic pharmacologic inhibition of SOCE ameliorated intestinal inflammation compared to controls, although the differences in histological colitis scores did not reach statistical significance at the end of the experiment on day 49 (**Figure 7C**). We observed significantly lower numbers of neutrophils in the colon Lamina propria (CLP) of CM4620 treated mice, whereas those of CD4^+^ T cells were not reduced, (**Figure 7D**). Further characterization of CD4^+^ T cell subsets in the LP revealed that treatment with CM4620 did not result in a decrease in the frequencies of RORγt^+^ Th17 cells and Foxp3^+^ Treg cells compared to control mice receiving vehicle only (**Figure 7E**). We next tested the function of CD4^+^ T cells isolated from the CLP and re-stimulated them *ex vivo* with PMA/ionomycin for 4h in the absence of CM4620. The frequencies of CD4^+^ T cells producing IFNγ, TNFα and IL-2 in the CLP of mice treated with SOCE inhibitor were significantly reduced compared to vehicle treated mice; moreover, cytokine levels per cell measured by MFI were significantly decreased (**Figure 7F**). Impaired IL-2 production was also observed after re-stimulation of CLP T cells with anti-CD3/CD28 for 24h (**Supplementary Figure 10**). Reduced production of IFN-γ, TNF-α and IL-2 is consistent with the suppressive effects of SOCE inhibition on CD4^+^ T cells amongst LPMC isolated from CD but especially UC patients (**Figure 4D**). Not all cytokines were affected by CM4620 treatment (at least after *ex vivo* re-stimulation of CLP derived T cells) because the frequencies of IL-17A^+^ IFN-γ^+^ producing CD4^+^ T cells were normal after P/I stimulation (**Supplementary Figure 10A**). Likewise, CD4^+^ T cells isolated from the CLP of CM4620 and vehicle treated mice produced similar amounts of IL-10 after re-stimulation with anti-CD3/CD28 for 24h *ex vivo* (**Supplementary Figure 10B**). It is noteworthy that serum levels of IFN-γ and TNF-α were unaltered in CM4620 treated animals compared to controls, whereas IL-6 levels were significantly decreased upon SOCE inhibition (**Figure 7G**). Collectively, our data indicate that SOCE inhibition after the onset of IBD is able to attenuate intestinal inflammation, which is mediated at least partially by suppression of pro-inflammatory cytokine production, while preserving the numbers of Treg cells in the CLP.

**Figure 7.**
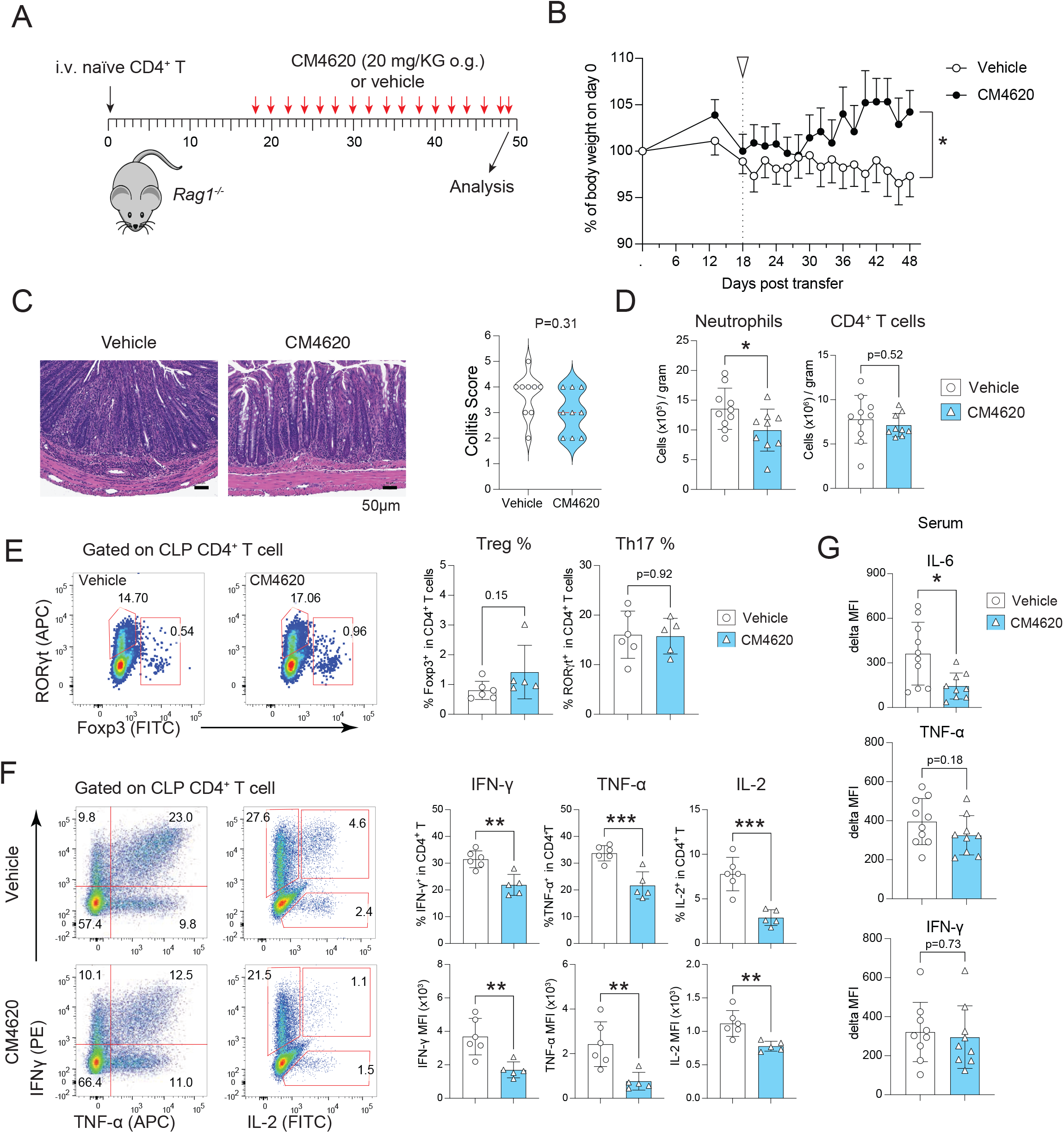
CRAC channel inhibitor CM4620 alleviates colon inflammation. (**A**) IBD was induced by i.v. injection of 5 x 10^5^ naïve CD4^+^ T cells into *Rag1^−/−^* host mice followed by oral gavage of mice with 20 mg/KG CM4620 or vehicle (SDD) every other day from days 18-49. (**B**) Relative weight loss of mice treated with vehicle or CM4620; the start of treatment is indicated by the white arrow. (**C**) Representative H&E staining of the distal colon of *Rag1^−/−^* mice treated with vehicle or CM4620. Colitis scores of 9 mice per cohort. Each symbol represents one mouse. Statistical analysis by Mann-Whitney U test. (**D**) Number of CD11b^+^ Gr-1^+^ neutrophils and CD4^+^ T cells (normalized to gram tissue) in the colon lamina propria (CLP) of mice on day 49 quantified by flow cytometry. Each dot represents one mouse. Data are the mean ± SEM of 9 mice per cohort. (**E**) Frequencies of RORγt^+^ Th17 cells and Foxp3^+^ Treg cells in the CLP of mice treated with CM4620 or vehicle. Each dot represents one mouse. Data are the mean ± SEM of 6 control and 5 CM4620 treated mice. (**F**) Frequencies of IFN-γ, TNF-α and IL-2 producing CD4^+^ T cells isolated from the CLP, restimulated *ex vivo* with PMA and ionomycin for 4 h and analyzed by flow cytometry. Bar graphs represent the percentages of IFN-γ^+^, TNF-α^+^, IL-2^+^ cells in CD4^+^ cells and mean fluorescent intensity (MFI) of IFN-γ, TNF-α, IL-2 on CD4^+^ cells, respectively. Each dot represents one mouse. Data are the mean ± SEM of 6 control and 5 CM4620 treated mice. (**G**) Levels of IL-6, IFN-γ and TNF-α in the serum of CM4620 and vehicle treated mice at day 49 measured by cytometry bead assay (CBA). Each dot represents one mouse. Data are the mean ± SEM of 10 control and 9 CM4620 treated mice. Statistical analyses in panels D-G by unpaired student’s t-test: ***p<0.001 **p<0.01 *p<0.05.

## Discussion

We here characterize the immune cell composition in the colon LP of patients with UC and CD by mass cytometry and show that SOCE regulates the function of immune cells driving IBD pathology. We also demonstrate that inhibition of SOCE, by genetic deletion of CRAC channel genes or pharmacological inhibition, attenuates intestinal inflammation in mouse models of colitis. By contrast, exposure to the SOCE inhibitor BTP2 does not affect the function and differentiation of primary murine and human intestinal epithelial cells. Together, our data demonstrate that SOCE is an important driver of the pro-inflammatory function of immune cells in IBD and suggest that inhibition of SOCE may represent a new approach for the treatment of CD and UC patients.

To characterize the immune cell composition in the colonic LP of therapy-refractory UC and CD patients compared with non-inflamed controls we used mass cytometry. Shared hallmarks of altered T cell composition in both IBD patient cohorts were an increased abundance of naive and effector CD4^+^ T cells and naive CD8^+^ T cells, whereas the frequencies of CD4^+^CD45RO^+^IL-7R^+^ effector memory T cells and CD8^+^ effector T cells were decreased. The overall frequencies of Treg cells were increased in the colonic LP of both UC and CD patients, but a significantly larger percentage of the remaining Treg cells produced TNFα and IL-17. These findings are in line with published single cell RNA sequencing data that also describe a higher expression of inflammatory cytokines in Treg cells of IBD patients (3), supporting the notion that intestinal inflammation in IBD is driven at least in part by an imbalance of effector versus regulatory T cells and altered Treg cell properties such as the production of inflammatory cytokines. Surprisingly, IL-4 and IL-13 producing Th2 cells were significantly reduced in both UC and CD, suggesting that Th2 cells may not be the main source of these cytokines in IBD, reminiscent of a predominant production of IL-4 and IL-13 by NKT cells in allergen-induced airway hyper-reactivity (33). In the B cell compartment, we observed a significant increase in the number of cells producing IL-6 and a reduction of CD38^+^IgA^+^ cells in both UC and CD patients. The depletion of B cells producing IgA, which is required for the opsonization and neutralization of pathogenic bacteria in the gut, is likely to contribute to IBD in both UC and CD patients, which is also supported by the fact that patients with common variable immunodeficiency and IgA deficiency are prone to develop IBD-like inflammatory enteropathies in 4-10% of cases (34, 35).

Besides these commonalities between both IBD patient cohorts, UC and CD patients also featured several entity-specific differences in their LPMC composition: Thus, the colonic LP of UC patients, but not CD patients, was infiltrated with PD1 expressing CD4^+^ and CD8^+^ effector T cells, CD4^+^ effector memory T cells and Treg cells, which points to a chronic activation of these T cells, potentially due to translocation of intestinal antigens into the LP. Expression of the inhibitory receptor PD1 on antigen activated T-cells in UC patients might thereby represent an attempt to maintain immune tolerance and to limit excessive T cell responses. Abu-Sbeih et al. recently reported a significantly higher rate of adverse gastrointestinal effects including diarrhea and colon perforation in 41% of IBD patients receiving checkpoint inhibitors compared to 11% in non-IBD patients (36). Our finding that the frequencies of PD1^+^ T cells are significantly increased in the colonic LP of UC patients provides a potential explanation for this clinical observation and cautions against the use of checkpoint inhibitors such as Nivolumab or Pembrolizumab in IBD patients because of their potential to induce severe flairs in UC patients. In line with the concept of a partially exhausted T cell phenotype due to chronic inflammation in UC patients, the production of the inflammatory cytokines IFNγ and IL-6 and the expression of the NFAT target CD103 (ITGAE) (37) were reduced both in effector and effector memory CD4^+^ T cells of UC patients, when compared to non-inflamed control samples and especially compared to CD patients. In contrast to UC patients, the immune cell landscape in the LP of CD patients was mainly characterized by high frequencies of CD38^+^ B cell clusters producing IgM or IgA as well as IFNγ-producing NK cells and IL7R-expressing CD4^+^ and CD8^+^ memory T cells, suggesting that these cells contribute to disease pathology in CD patients.

The production of cytokines by immune cells in the LP is an important pathogenic driver of intestinal inflammation in UC and CD. In CD, IFNγ and IL-17/IL-22 generated by Th1 and Th17 cells were early on described as dominant cytokines potentially involved in disease pathogenesis, whereas the development of UC has been classically linked to high levels of IL-4 and IL-13 produced by T cells and NKT cells (38). Remarkably, our mass cytometric data did not find a predominance of IL-4 and IL-13 in the LP of UC patients but we instead mainly detected cells producing the pro-inflammatory cytokines IFNγ, TNFα and IL-6 that are known to be present in both forms of IBD (38). Intriguingly, we did not observe significant differences in the expression levels of pro-inflammatory cytokines on a single cell basis in LPMC isolated from surgical colon specimen of CD and UC patients when compared to non-inflamed controls, but we observed a higher frequency of cells expressing pro-inflammatory cytokines instead. However, we used a strong stimulus (PMA plus ionomycin) to induce cytokine production, which may also mask differences in cytokine production by LPMC in inflamed and non-inflamed regions. Only Treg cells isolated from inflamed lesions of CD and UC patients showed significantly increased expression of IL-17 and TNFα relative to Treg cells from non-inflamed control patients.

The expression of the inflammatory cytokines described above is regulated by Ca^2+^ dependent transcription factors including most prominently NFAT (10). Given the important role of these cytokines in IBD pathogenesis, we investigated how inhibition of SOCE as the main Ca^2+^ influx pathway in immune cells affects cytokine production. Using the CRAC channel inhibitor BTP2 we here demonstrate that SOCE is required for the production of many inflammatory cytokines including IL-2, IL-4, IL-17, TNFα and INFγ by T cells and, although to a lesser degree, B cells and myeloid cells. Increasing concentrations of BTP2 resulted in a dose-dependent suppression of cytokine production in CD4^+^ and CD8^+^ effector and effector memory T cells, Th17 cells as well as Treg cells isolated from the LP of IBD patients. The most Ca^2+^ dependent cytokines were IL-2 and IFNγ, followed by TNFα and IL-17, whereas the Th2 cytokines IL-4 and IL-13 were surprisingly insensitive to BTP2-mediated SOCE inhibition. When we compared the effects of BTP2 on cytokine production by a variety of colonic T cell subsets, we observed significant reductions in the expression of IL-2, TNFα and IFNγ in both CD and UC patients. A more selective suppression of IL-4 and IL-13 production by BTP2 was found in UC patients. IL-17 production was significantly suppressed by BTP2 only in T cells of CD patients and interestingly only in CD4^+^ T cell subsets that also expressed TNFα. The suppressive effects of SOCE inhibition on cytokine expression in T cells of IBD patients are consistent with a report of decreased production of IFNγ, IL-2 and IL-17 by LPMCs of IBD patients that had been stimulated *ex vivo* in the presence of the SOCE inhibitor Synta 66 (39). Significant suppression of cytokine production by BTP2 was also observed for TNFα, IFNγ and IL-2 in Treg and NK cells of CD patients, and for TNFα, IL-2 and IL-17 in ILCs of UC patients. BTP2 also blocked the production of TNFα, IL-2 and IL-6 by B cells of both UC and CD patients, whereas the effects of SOCE inhibition in myeloid cells were limited to reduced IFNγ production in CD patients. These broad effects of SOCE inhibition on the production of pro-inflammatory cytokines by T, B and myeloid cells of UC and CD patients emphasize the important role of SOCE in the transcriptional regulation of these cytokines and the potential of CRAC channel blockers for the treatment of IBD.

To investigate whether the suppressive effects of SOCE inhibition on the *ex vivo* production of pro-inflammatory cytokines by LPMCs isolated from UC and CD patients translate into attenuated IBD severity *in vivo*, we used a murine T cell transfer model of IBD. Whereas naïve CD4^+^ T cells of wild-type mice induced colitis in lymphopenic host mice, deletion of *Stim1* in CD4^+^ T cells and, to a lesser degree, *Orai1* attenuated IBD severity, whereas deletion of *Stim2* had no significant protective effect. The degree of IBD attenuation correlated with the degree of reduced production of IFNγ, IL-17A and TNFα by T cells isolated from the mLN of mice. The deletion of *Stim2*, Orai1, *Stim1* and *Stim1/Stim2* together in T cells results in progressively more pronounced defects in SOCE and, consequently, the expression of IFNγ by Th1 cells and IL-17A by Th17 cells *in vitro*. Importantly, the production of IFNγ and IL-17A was almost abolished in *Orai1-*deficient T cells despite substantial residual SOCE, demonstrating that incomplete suppression of SOCE is sufficient to strongly attenuate inflammatory cytokine production. By contrast, the differentiation of induced Foxp3^+^ iTreg cells *in vitro* was less affected in *Orai1*-deficient T cells. Only *Stim1-* or *Stim1/Stim2*-deficient T cells with strongly reduced or abolished SOCE displayed severe defects in the differentiation of iTreg cells *in vitro*. These data indicate that effector Th1 and Th17 cells require strong Ca^2+^ influx for the production of pro-inflammatory cytokines, whereas moderate SOCE signaling strength is sufficient for the differentiation of tolerogenic iTreg cells. These findings are consistent with the attenuated severity of IBD after adoptive transfer of ORAI1- or STIM1-deficient CD4^+^ T cells. Genetic deletion of *Orai1* and partial suppression of SOCE in T cells also attenuated IFNγ and IL-17 production and the severity of EAE, a mouse model of the autoimmune disease multiple sclerosis, in which Th1 and Th17 cells play important pathogenic roles (21, 22, 24, 26). By contrast, the development and suppressive function of nTreg and iTreg cells are normal in mice with conditional deletion of *Orai1* (26, 40) or *Stim1* (29). These data suggest that a therapeutic window exists for the inhibition of pro-inflammatory effector T cells without compromising the tolerogenic function of Treg cells. The concept of different SOCE thresholds in pro-inflammatory effector and regulatory T cells may extent to human immune cells. Thus, we observed that the production of IFNγ, IL-2 and other cytokines was reduced by > 50% using BTP2 in the sub-micromolar range in CD4^+^ effector and memory T cells of CD and UC patients. By contrast, a concentration of 1 μM BTP2 was required for a > 50% reduction of IL-17 and TNFα production by Treg cells. Moreover, cytokine production by B cells and myeloid cells was not or only moderately affected by SOCE inhibition even after exposure to high concentrations of BTP2. The molecular mechanisms controlling these differential SOCE requirements in pro-inflammatory T cells versus other immune cell subsets require further study as they might offer the opportunity to selectively control the function of specific immune cell subsets. Collectively, our data suggest that SOCE inhibition might represent an effective treatment of IBD due its preferential effects on the pro-inflammatory function of T cells.

To investigate whether pharmacologic inhibition of CRAC channel function is an effective and safe approach to IBD treatment, we tested the effects of a systemic application of the CRAC channel inhibitor CM4620 in a murine adoptive T cell transfer model of IBD. At serum concentrations equivalent to the IC_50_ for inhibition of human T cell function *in vitro*, we observed reduced weight loss, neutrophil inflammation of the LP and production of IFNγ and TNFα by LP T cells, whereas the frequencies of Treg cells and the amount of secreted IL-10 were not altered in the LP of animals after blockade of SOCE. These results indicate that SOCE inhibition attenuates IBD severity even when treatment is started after disease initiation and administered systemically. This is significant given the fact that SOCE is the source of Ca^2+^ signals in many non-excitable cell types, including intestinal epithelial cells. To exclude that CRAC channel inhibition has adverse effects on IEC function and the intestinal barrier, thereby exacerbating IBD, we treated primary murine and human colonic epithelial cells grown in organoid cultures with the selective CRAC channel inhibitor BTP2 *in vitro.* Exposure to 1 µM BTP2 over 4 days did not affect the survival and differentiation of IECs or trans-epithelial resistance suggesting that SOCE inhibition does not significantly impair IEC and intestinal barrier function. We cannot rule out that SOCE inhibition may be required for the differentiation or function of rare epithelial cell subsets such as tuft cells, goblet cells or neuroendocrine cells, which play important roles in mucosal immune homeostasis, chemo-sensing and tissue regeneration (41, 42). Possible side effects of CRAC channel inhibitors on IEC and intestinal barrier function may manifest only after long-term application, which would be required for the treatment of IBD. Arguing against such a role of SOCE is the fact that patients with loss-of-function mutations in *ORAI1* or *STIM1* genes and knock-out or knock-in mice lacking functional ORAI1 channels do not show signs of intestinal inflammation under steady state conditions (20, 43, 44). Patients with UC and CD involving the colon are at higher risk for developing colorectal cancer than the general population (45). A recent study reported that the phosphatase calcineurin and the transcription factor NFAT, which are activated by SOCE, contribute to the development of intestinal epithelial tumors by promoting the survival and proliferation of cancer stem cells (46). Inhibition of SOCE might therefore provide additional benefits to IBD patients by not only attenuating gastrointestinal inflammation but also by suppressing the growth of cancer stem cells.

The SOCE inhibitor CM4620 is currently tested in clinical trials as a novel treatment option for acute pancreatitis (NCT04195347) and COVID-19 pneumonia (NCT04345614). In patients with acute pancreatitis, CM4620 attenuated inflammation as evident from reduced pancreatic edema, acinar cell vacuolization, intra-pancreatic trypsin activity, cell death signaling and acinar cell death. CM4620 also decreased suppressed neutrophil functions and cytokine production by human peripheral blood mononuclear cells (45). In a randomized, controlled, open-label study in patients suffering from COVID-19-associated pneumonia, CM4620 treatment improved outcomes in patients with severe COVID-19 and had a favorable safety profile (47). These data suggest that SOCE inhibition is well tolerated and effective in patients with severe pulmonary and pancreatic inflammation. Both studies were limited by the small numbers of the patients treated with the CRAC channel inhibitor, and by the relatively short duration of treatment. Therefore, the efficacy and safety of long-term, systemic CRAC channel inhibition in chronic inflammatory diseases such as CD and UC remains to be studied.

## Data Availability

All mass and flow cytometric data sets are available from the corresponding authors on reasonable request that does not include confidential patient information.

## Acknowledgement

We would like to acknowledge the assistance of the BIH Cytometry Core (BIH and Charité – Universitätsmedizin Berlin, Germany) and the IBDome researchers Chiara Romagnani, Nils Müller, Ahmed Hegazy, Roodline Cineus, Julia Hecker, Nadine Sommer and Anja Kühl for supporting the isolation and processing of surgical specimen. This work was funded by the German Research Foundation (We 5303/3-1 to C.W., Si 749/10-1 to B.S., SFB-TRR 241 B01 to B.S. and C.W., SFB TRR167 B05 to CB) and NIH grants AI097302, AI130143 and AI137004 (to S.F.), a Senior Research Award by the Crohn’s and Colitis Foundation of America (to S.F.) and an Irma T. Hirschl career scientist award (to S.F.) M.B. and J.F.Z. received financial support from the Einstein Foundation Berlin. C.W. received funding by the Clinician Scientist Program of the Berlin Institute of Health and by the Fritz-Thyssen-Foundation (10.19.2.028MN).

## Author contributions

M.L., U.K., Y.-H.W., L.G., A.S., M.B., J.F.Z. and D.K. performed experiments. M.L., U.K., Y.-H.W., L.G., A.S., M.B., C.F.Z., C.B., S.S., K.S., D.K., S.F. and C.W designed experiments, analyzed and interpreted the data. B.S. contributed to clinical sample acquisition and helped with data interpretation. M.L., S.F. and C.W. wrote the paper.

## Online Materials and Methods

### Ethical regulations

Written informed consent was obtained from all patients and healthy donors including the consent to publish clinical information potentially identifying individuals. All experiments involving human material were approved by the institutional review board of the Charité-Universitätsmedizin Berlin and conducted accordingly. All animal experiments were performed in accordance to institutional guidelines for animal welfare approved by the Institutional Animal Care and Use Committee at New York University School of Medicine.

### Isolation of human lamina propria and peripheral blood mononuclear cells

**Surgical** colon resectates were obtained from 9 UC and 11 CD patients undergoing colectomy while non-inflamed specimen of 4 patients with colon rectal cancer served as non-inflamed controls (**Supplementary Table 1**). Colonic mucosa was detached using sterile forceps and washed in 1 mM 1,4 Dithiothret (DTT) (Roth) dissolved in Hanks’ Balanced Salt solution (HBSS) w/o Ca^2+^ and Mg^2+^ (ThermoFisher Scientific), in shaking conditions at RT for 15 min. Lamina propria was cleaned from any left submucosal tissue, cut in pieces and washed 3 times in 1 mM ethylenediaminetetraacetic acid (EDTA) (Sigma-Aldrich) under constant shaking for 15 minutes at 37°C in order to detach epithelial cells. Afterwards, EDTA was carefully washed out using HBSS w/o Ca^2+^ and Mg2^+^ and tissue was subsequently incubated in 0.15mg/ml collagenase A (ROCHE) for 16 h at 37°C under constant shaking. The supernatant was then filtered through a 100 µm cell strainer (ThermoFisher scientific) and washed three times in HBSS w/o Ca^2+^ and Mg^2+^. Cells were separated through a Percoll gradient (GE Healthcare) centrifugation for 30 min at 1500 rpm at 4°C. Lympho-monocytes were collected at the interface of 40-60% Percoll. Cells were washed twice and stimulated *ex-vivo* for 4 h in RPMI 1640 (ThermoFisher Scientific) supplemented with 10% fetal bovine serum (FBS; Sigma) and 10% Penicillin-Streptomycin (ThermoFisher Scientific) containing 20 ng/ml phorbol 12-myristate 13-acetate (PMA; Sigma-Aldrich) and 1 µg/ml ionomycin (Sigma-Aldrich) in the presence or absence of 0-1000nM BTP2 (Sigma Aldrich). Brefeldin A (10 µg/ml; Sigma-Aldrich) was added for 2 h, while 25KU Benzonase Nuclease (Sigma) was added (1:10.000) 15 min prior to harvesting. For mass cytometry, cells were fixed in Smart Tube buffer (SMART TUBE Inc.) supplemented with 20% BSA (ThermoFisher Scientific) and subsequently stored at −80°C until antibody staining.

### Calcium influx measurements in murine T cells

96-well-imaging plates (Fisher) were coated with 0.01% poly-L-lysine (w/v) (Sigma-Aldrich) diluted in water for 2 h and then washed with sterile water. Cells were labeled with 2 µM Fura-2 AM (Life Technologies) for 30 min in cell culture medium and attached for 10 min to the plates. Intracellular Ca^2+^ measurements were analyzed using a Flexstation 3 fluorescence plate reader (Molecular Devices) at 340 and 380 nm excitation wavelengths. Cells were stimulated with 0.3 µM ionomycin (EMD Millipore) in 1 mM Ca^2+^ Ringer solution (155 mM NaCl, 4.5 mM KCl, 3 mM MgCl2, 10 mM D-glucose, 5 mM Na HEPES) to induce SOCE. Fura-2 fluorescence emission ratios (F340/380) were collected at 510 nm following every 5 s. Ca^2+^ signals were quantified by analyzing the peak value of F340/380 ratios using the GraphPad Prism 6.0 software.

### Calcium influx measurements in human LPMCs and PBMCs

Heparinized blood was obtained from three healthy donors and PBMCs were isolated by density gradient centrifugation using Biocoll (Merck). Alternatively, surgical colon specimen were obtained from three IBD patients undergoing colectomy and cells were isolated according to the isolation of lamina propria mononuclear cells protocol described above. Isolated PBMCs or LPMCs were cultured for 16h at 37°C, 5% CO_2_ in the presence or absence of 15 – 1,000 nM BTP-2 (Sigma Aldrich) dissolved in dimethyl sulfoxide (DMSO) (Sigma). Subsequently, cells were loaded with the calcium sensing dye Fluo-4 AM/DMSO (2µg/ml; Life Technology) for 30 min on ice protected from light, washed with PBS/5% BSA (ThermoFisher Scientific) and afterwards stained for 15 min on ice with anti-human CD3 (APC, OKT1, eBioscience) or CD3 (Viogreen, REA613, Miltenyi Biotec), CD4 (BV510, RPA-T4, Biolegend), CD8 (APC-Cy7, SK1, Biolegend), CD19 (APC-Cy7, HIB-19, Biolegend), CD14 (APC, 63D3, Biolegend), CD16 (Pe-Cy7, 3G8, BD) and CD56 (APC, TULY56, eBioscience). To measure baseline intracellular Ca^2+^ ([Ca^2+]^i) cells were washed and re-suspended in 0mM Ca^2+^ Ringer solution (155 mM NaCl, 4.5 mM KCl, 3 mM MgCl_2_, 10 mM D-glucose, 5 mM Na-HEPES) and acquired for 30 sec using a Canto II flow cytometer (BD Bioscience). Subsequently, cells were stimulated with 1 µM thapsigargin (EMD Millipore, Billerica, MA), Ca^2+^ containing Ringer solution (2 mM CaCl_2_) was added after 300 sec to the cells (final [Ca^2+^]o 1 mM). The sample was subsequently acquired for additional 120 sec. For the analysis, the mean fluorescence intensity (MFI) of Fluo-4 (*f*) was normalized to the MFI average detected during the 30s baseline measurement (*f_0_*) and the resulting ratio *f/f_0_* was plotted against a time (t) axis. Graphs were plotted using Prism 8 software.

### Annexin V and 7AAD staining of human LPMCs

Colon resectates were obtained from 3 IBD patients undergoing colectomy and cells were isolated according to the isolation of lamina propria mononuclear cells protocol described above. Cells were stimulated *ex-vivo* for 4 h in RPMI 1640 (ThermoFisher Scientific) supplemented with 10% fetal bovine serum (FBS; Sigma) and 10% Penicillin-Streptomycin (ThermoFisher Scientific) containing 20 ng/ml phorbol 12-myristate 13-acetate (PMA; Sigma-Aldrich) and 1 µg/ml ionomycin (Sigma-Aldrich) in the presence or absence of 1µM BTP2 (Sigma Aldrich). Brefeldin A (10 µg/ml; Sigma-Aldrich) was added 2h prior to harvesting. Cells were stained according to Pacific Blue™ Annexin V Apoptosis Detection Kit with 7-AAD (Biolegend). Samples were measured using a Canto II flow cytometer (BD Bioscience). Data were analyzed using the FlowJo software package V10.1 (FlowJo, LLC).

### Mass cytometry staining and acquisition

Cells were thawed and barcoded for 30 min at RT by using the Cell-ID 20-plex Pd Barcoding Kit (Fluidigm) ± CD45-89Y staining in order to pool more samples in one batch/run (max 40 samples). Individual samples were washed twice with cell staining buffer (Fluidigm) and pooled together before staining. Anti-human antibodies were purchased either pre-conjugated to metal isotopes or conjugated in house using the conjugation MaxPar X8 kit (Fluidigm) according to the manufacturer’s protocol (**Supplementary Table 2**). For surface staining, cells were incubated with antibodies for 30 min at 4 °C, then washed twice with cell staining buffer and incubated for 60 min at 4 °C in fixation/permeabilization buffer according to the manufacturer’s protocol (Fix/Perm Buffer, eBioscience). Cells were then washed twice using permeabilization buffer (eBioscience) and stained with antibody cocktails against intracellular molecules for 1h at 4°C. Cells were subsequently washed twice with permeabilization buffer and incubated overnight in 2% methanol-free formaldehyde solution (ThermoFisher). Cells were washed and re-suspended in 1 ml iridium intercalator solution (Fluidigm) for 1 h at RT. Finally, cells were washed twice with cell staining buffer and then twice with Maxpar Water (Fluidigm) for CyTOF measurements. Cells were analyzed using a CyTOF2 upgraded to Helios specifications, with software version 6.5.236, as previously described (48). Compensation beads staining: OneComp eBeads™ Compensation Beads (Invitrogen) were stained individually with each of the antibodies included in our panel (**Supplementary Table 2**) for bead-based compensation of mass cytometry. For each channel assessed in the panel described above, one drop of OneComp eBeads was loaded in a well of a v-bottom 96 well plate (Corning) and individually stained with 1 µg of the corresponding metal-labeled antibody for 30 min at room temperature. After staining, individual beads were washed three times in staining buffer and afterwards pooled in a single tube and washed once in PBS (ThermoFisher Scientific). Beads were then fixed in 1.6% methanol-free formaldehyde solution (ThermoFisher) for 1 h at room temperature. After fixation, beads were washed twice in staining buffer and twice in Maxpar water. Beads were re-suspended in 300 µL of Maxpar water previous CyTOF acquisition.

### Data analysis of mass cytometry data

The Cytobank software package (www.cytobank.org) was used for initial manual gating on single cells and for de-barcoding of pooled samples. FCS files of de-barcoded single-cells were exported and further compensated for signal spillover using the R Software (3.6.0, Bioconductor 9) and by applying the *CATALYST* package and arcsinh transformation (scale factor 5) prior to data analysis. Compensated files were then gated on CD45^+^, CD45^+^CD3^+^ or CD45^+^CD3^−^ cells and t-SNE maps of each pre-gated cell population were generated according to the expression of markers listed in **Supplementary Table 3-6** by using the Cytobank Software. Subsequently, FCS files containing the t-SNE embedding as additional two parameters were exported from Cytobank and the clustering analysis was performed using the *FlowSOM/ConsensusClusterPlus* package on R (3.6.0, Bioconductor 9) (**Supplementary Fig. 7**). Cell clusters of LPMCs were identified after visual inspection of t-SNE plots and functional interpretation of heat maps generated by *FlowSOM/ConsensusClusterPlus*. Statistical significance of differential abundant clusters (DA) among the 3 disease groups (UC, CD and Non-inflamed) was performed using a generalized linear mixed-effects model (GLMM) available through the R package *diffcyt* (used all defaults with analysis_type  =  “DA”, method_DA  =  “diffcyt-DA-GLMM”, min-cells  =  3). False discovery rate (FDR) was adjusted at 10% using the Benjamini-Hochberg (BH) procedure for multiple hypothesis testing, as previously described (25). GraphPad Prims 8 and multiple t test (two-stage linear step-up procedure of Benjamini, Krieger and Yekutieli, with Q = 1%) were applied in order to analyze protein expression levels among 3 disease groups (UC, CD and Non-inflamed), while the effects of BTP2 on LPMCs isolated from UC or CD patients were calculated by a paired Wilcoxon matched-pairs signed rank test, *p < 0,05.

### Human and murine epithelial cells acquisition

Intestinal tissue was obtained from colon resectates of two CD patients and colon crypts were isolated from non-inflamed areas of the ascending colon. Intestinal tissue was washed and cut into smaller pieces (approx. 3 mm) after removing underlying muscle layers with surgical scissors. For murine samples, *C57BL/6* mice between 8 and 15 weeks were euthanized and the colonic part of the gut was removed.

### Crypt Isolation and culture

Establishment and maintenance of murine and human spheroid cultures from the colon epithelium were obtained as described previously (49–51). In brief, colons from C57B/6 wild-type mice were opened longitudinally, cut into small pieces, and washed with cold phosphate-buffered saline (PBS; Gibco; Life Technologies, Carlsbad, USA). Human intestinal mucosa was isolated by removing the muscle and fat tissue from colon resectates. The tissue was then cut into small (1-2 mm) stripes and washed with cold PBS. Intestinal fragments were washed repeatedly with cold PBS until the supernatant was clear. The tissue was then incubated with 10 mM EDTA cold chelation buffer (PBS supplemented with 54.9 mM D-sorbitol, 43.4 mM sucrose, and 1 mM 1,4-dithiothreitol (DTT; Carl Roth, Karlsruhe, Germany) and an antibiotic cocktail containing ciprofloxacin (10µg/ml; Fresenius Kabi, Bad Homburg, Germany), fluconazol (1µg/ml; B. Braun, Melsungen, Germany), primocin (100µg/ml; InvivoGen, San Diego, USA) and gentamycin (50µg/ml, Ratiofarm, Ulm, Germany)) for 45 minutes on ice with gentle shaking. EDTA-containing supernatant was removed, 10ml chelation buffer (without DTT and antibiotics) was added and crypts were released from the tissue by vortexing four times for 30 seconds. Supernatant of every vortexing step was pooled and washed (300g, 4°C, 5 minutes) two times with 10 ml washing medium (advanced DMEM/F-12 (Gibco, Life Technologies, Carlsbad, USA) supplemented with 2 mM L-glutamine, 50 units/ml penicillin, and 50 µg/ml streptomycin, 10 mM HEPES (Gibco). Crypts were transferred to a 1.5-ml microtube followed by seeding.

Isolated intestinal crypts were counted, embedded in Matrigel (Corning, NY, USA; growth factor reduced, phenol red-free, LDEV-free) on ice and seeded into 24-well plates (500 crypts in 50 µl Matrigel per well). Matrigel was polymerized for 15 minutes at 37°C and overlaid with 500 µl of spheroid culture medium (50% L-WRN-CM2 [conditioned medium produced using the cell line L-WRN CRL-3276, ATCC, Manassas, USA] in advanced DMEM/F-12 supplemented with 20% fetal bovine serum, 2 mM L-glutamine, 50 units/ml penicillin, and 50 µg/ml streptomycin (Merck Millipore, Burlington, USA) (and additional supplements specific for mouse and human [Mouse: 50 ng/ml recombinant murine EGF (Peprotech, Hamburg, Germany). Human: 50 ng/ml recombinant human EGF (Peprotech), 10 mM nicotinamide (Sigma, St. Louis, USA), 0.5 µM A83-01, 10 µM SB202190, 10 nM human gastrin I] (all from Sigma)). 10 µM Y-27632 (Abmole Bioscience, Houston, USA) was added to the culture medium for the first two days of culture after isolation or passaging. Fresh culture medium was added every other day and spheroids were passaged once a week at a 1:6 splitting ratio. For passaging, Matrigel was scraped off to collect organoids in a 15-ml conical tube. Organoids were digested into single cells in 1 ml TrypLE (Gibco) for 5 minutes at 37°C, embedded in Matrigel and seeded into a 24-well plate.

### Resazurin Viability Assay

Spheroids were passaged, embedded in Matrigel and seeded into 96-well plates at a density of approx. 100 organoids in 8 µl Matrigel per well. Organoids were cultured in spheroid culture medium for 5 days. 10 µM Y-27632 was added for the first two days of culture. To induce differentiation, WRN-CM, nicotinamide, and SB202190 were withdrawn from the medium and 100 ng/ml recombinant noggin (Peprotech), 500 ng/ml recombinant human R-Spondin (R&D Systems, Wiesbaden, Germany) and 5 µM DAPT (Selleckchem, Houston, USA) were added as described previously (49). Initial organoid viability was assessed with resazurin sodium salt (Sigma) once before changing to differentiation medium. Resazurin was added to every well at a working concentration of 0.25 µg/ml and the culture was incubated in the dark at 37°C for one hour. Subsequently, the culture medium was transferred to a black flat clear-bottom 96-well plate and fluorescence was measured at 560 nm excitation and 590 nm emission with a SpectraMax Gemini EM Microplate Reader (Molecular Devices, San Jose, USA). Wells with organoid culture were washed with PBS and fresh medium was added. Culture was continued at 37°C and viability was measured with resazurin as described above on the indicated days of culture. The medium was supplemented with 1 µM BTP2 starting from day 2 of differentiation.

### Establishment of 2D-Monolayers and Treatment with BTP2 and TNFα and IFNγ

Organoid-derived monolayers were generated based on published protocols (52, 53). Briefly, the upper compartment of a 0.6 cm^2^ transwell filter (Corning; PCF; 0.4 µm pores) was coated with 150µl Matrigel mixed 1:10 with Advanced DMEM/F-12 after chilling the plate at −20°C for 30 min and incubated for 16 h at 4°C. Before seeding, coating solution was removed and the plate was warmed at 37°C for 30 minutes. After 5 days of culture, spheroids were digested into single cells and seeded at approx. 0.3 x 10^6^ cells per filter in 400 µl pre-warmed (37°C) spheroid culture medium supplemented with ROCK1 inhibitor Y-27632 for the first two days of culture. Transepithelial electrical resistance (TEER) was measured with an ohmmeter (Millicell® ERS-2 Volt-Ohm Meter; Merck Millipore). Organoids were cultured until a stable TEER was reached, differentiation was induced as described above and BTP2 (1µM), TNF-α (10ng/ml) and IFNγ (10ng/ml) were added as indicated.

### Mice

*Stim1^fl/fl^-CD4-Cre*, *Stim2^fl/fl^-CD4-Cre* (*21*), *Stim1^fl/fl^Stim2^fl/fl^-CD4-Cre* (54) or *Orai1^fl/fl^-CD4-Cre* mice (26) have been described previously. *Rag1^−/−^* mice were purchased from The Jackson Laboratory (stock number 002216). Animals were used between 6 and 12 weeks of age. Both female and male mice were included in the studies and no mice were excluded. All mice were maintained under specific pathogen-free conditions.

### In vitro activation and differentiation of murine T cells

Naïve T cells were isolated from spleens and LNs of *Stim1^fl/fl^-CD4-Cre*, *Stim2^fl/fl-^CD4-Cre*, *Stim1^fl/fl^ Stim^2fl/fl-^CD4-Cre* or *Orai1^fl/fl^-CD4-Cre* mice and stimulated with 1 µg/ml plate-bound anti-CD3 (clone 2C11 obtained from BioXCell) and 1 µg/ml anti-CD28 (clone 37.51,obtained from BioXCell) for 3 days in the presence of the following cytokine cocktails for differentiation into Th1: 10 ng/ml IL-12 (PeproTech) and 2 µg/ml anti-IL-4 (eBioscience), Th17: 20 ng/ml IL-6 (PeproTech), 0.5 ng/ml human TGF-β1 (PeproTech), 2 µg/ml anti-IL-4 and 2 µg/ml anti-IFNγ (eBioscience), or iTreg: 2.5 ng/ml TGF-β1 (PeproTech) in IMDM (Cellgro) containing 2 mM L-glutamine, 50 µM 2-Mercaptoethanol, 100 U/ml penicillin, 100 µg/ml streptomycin and 10% FCS.

### T cell transfer colitis

Naïve CD4^+^ T cells were isolated from *Stim1^fl/fl^-CD4-Cre*, *Stim2^fl/fl-^CD4-Cre*, *Stim1^fl/fl^ Stim^2fl/fl-^CD4-Cre* or *Orai1^fl/fl^-CD4-Cre* mice by flow cytometric cell sorting for CD4^+^ CD62L^+^ CD25^−^ CD45RB^hi^ using a SONY SY3200 cell sorter. 5 x 10^5^ cells were subsequently injected intra-peritoneally into recipient *Rag1^−/−^* mice. After adoptive transfer of T cells, *Rag1^−/−^* mice were monitored weekly for body weight loss over a period of 8 weeks.

### T cell transfer colitis and in vivo treatment with CM4620

Naïve CD4^+^CD44^low^CD62L^+^CD25^−^ T cells were FACS sorted and transferred via intravenous (i.v.) injection into sex matched *Rag1^−/−^* mice (5×10^5^ cells per mouse) and weight loss was monitored. 18 days after T cell transfer, mice were either treated with the selective CRAC channel inhibitor CM4620 (kindly provided by CalciMedica, La Jolla, CA) or treated with vehicle controls (55). The CM4620 compound was reconstituted in a vehicle composed of 0.5% methylcellulose (w/w) with a viscosity of 400 cP (Sigma Aldrich, M0262) and 1% w/w Tween 80 (Company, P1754) in ddH2O. The active form of CM4620 was administered by oral gavage at 20 mg per kg bodyweight (25% loading) plus 75 mg/kg bodyweight of hypromellose acetate succinate (HPMCAS) bead carrier formulated in vehicle every other day until the end point. Mice were sacrificed 7 weeks post T cell transfer. Samples of distal and proximal colon were fixed with 4% of paraformaldehyde. Paraffin-embedded samples were cut into 5 µm sections both hematoxylin and eosin staining as well as alcian blue staining were performed. Colon inflammation was scored by a double blinded pathologist described above.

### Histopathologic scoring of intestinal inflammation in mice

8 weeks after induction of T cell transfer colitis, colon sections were fixed in 4% paraformaldehyde, embedded in paraffin and stained with haematoxylin and eosin (H&E) using standard protocols. Histology sections were scored by two independent researchers in a blinded fashion using the following grading system: 0: no changes; 1: minimal scattered mucosal inflammatory cell infiltrates, with or without epithelial hyperplasia; 2: mild scattered to diffuse inflammatory cell infiltrates, sometimes extending to the submucosa and associated with erosions, with mild epithelial hyperplasia and minimal to mild mucin depletion from goblet cells; 3: mild to moderate inflammatory cell infiltrates, sometimes trans-mural, often associated with ulceration, with moderate epithelial hyperplasia and mucin depletion; 4: marked inflammatory cell infiltrates that were often transmural and associated with ulceration, with marked epithelial hyperplasia and mucin depletion; 5: marked transmural inflammation with severe ulceration and loss of intestinal glands.

### Isolation of murine Lamina propria leucocytes

Colon tissue of mice was opened longitudinally by a curved scissor and cut into pieces around 0.5 to 1cm, colon bits were predigested with pre-digestion buffer (5mM EDTA and 1mM DTT in HBSS buffer without calcium) for 15 minutes. Colon bits were then washed, further minced and digested with digestion buffer (1mg/mL collagenase IV and 0.1mg/mL DNase-I in HBSS with 5% FBS) in 37C shaker at a speed of 225rpm for 45 minutes. Cells were then isolated using a 40/75% discontinuous Percoll gradient, the middle layer was collected, washed, and re-suspended in RPMI1640 with 2% of fetal bovine serum. Cells were counted using hemocytometer.

### Flow cytometric analyses of murine LPMCs from CM4620 treated mice

For surface antigen staining, cells were prepared in PBS containing 2% of FBS and 2 mM EDTA. Cells were stained with fluorescently labeled antibodies for 20 min on ice in the dark. For intracellular cytokine detection, leucocytes were cultured in RPMI1640 plus 10% FBS with the stimulation with 20 nM phorbol 12-myristate 13-acetate (PMA) (PMA, Calbiochem, 524400) and 0.5 µM ionomycin (Invitrogen, I-24222) with Golgi stop 5 µM brefeldin A (eBioscience, 00-4506-51) for 4 h, followed by washing the cells in PBS 2% FBS and blocked with anti-CD16/CD32 antibodies (2.4G2, BioXcell) for 15 minutes. Surface staining was performed using molecules with fluorescently labeled antibodies for 20 min on ice in the dark. For intracellular cytokine staining, cells were fixed with IC Fixation Buffer (eBioscience, Cat: 00-8222-49) for 30 minutes and permeabilizated using Permeabilization Buffer (eBioscience, Cat: 00-8333-56) following washing and intracellularly staining procedure. FACS samples were analyzed on a BD LSR II Fortessa cytometer and data were analyzed using the FlowJo software version 10.6.2. A list of fluorescent conjugated antibodies can be found in **Supplementary Table 7.**

### Cytometry bead assay

For measuring the cytokine production by T cells in colon, colon lamina propria derived total lymphocytes were stimulated with 0.5ug/mL anti-CD3 (clone 145-2C11, BioXcell) for 24 hours, supernatant was collected and cytokine levels were measured using a cytometry bead assay (BD Bioscience). Cytokines levels in serum from host mice were also measured using a Th1/Th2/Th17 BCA Kit (Cat 560485, BD Bioscience) as instructed by vendor. Positive and negative controls were analyzed simultaneously.

