## Supplementary data for "Store-Operated Calcium Entry Controls Innate and Adaptive Immune Cell Function in Inflammatory Bowel Disease"

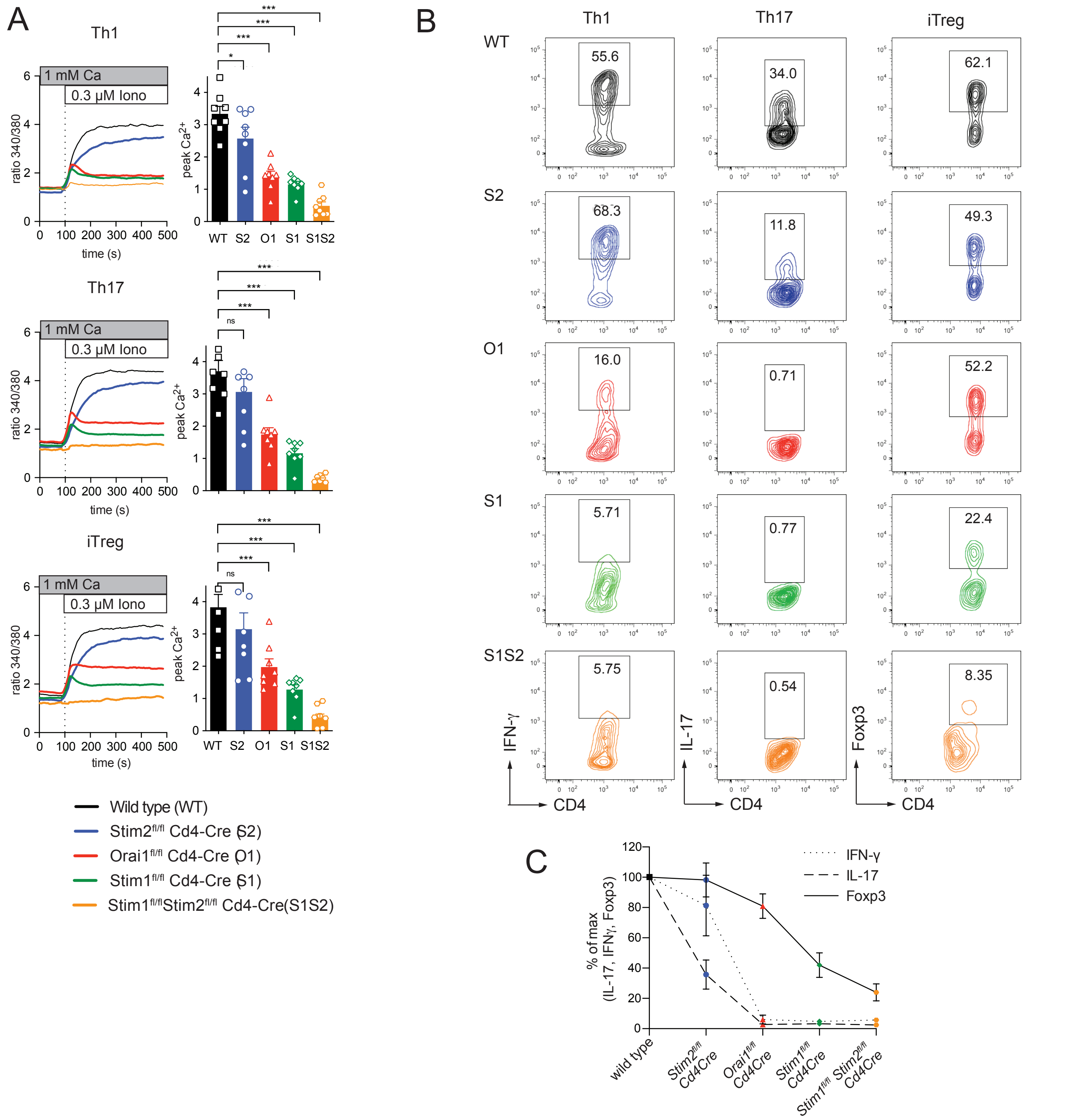

**Supplementary Figure 1**. Distinct quantitative SOCE requirements in CD4+ T cell subsets. Naïve CD4+ T cells were isolated from spleen and LNs of *WT*, *Stim2fl/fl-Cd4-Cre (S2)*, *Orai1fl/fl-Cd4-Cre (O1)*, *Stim1fl/fl-Cd4-Cre (S1)* and *Stim1flflStim2fl/fl-Cd4-Cre* (S1S2) deficient mice and differentiated into Th1, Th17 and iTreg cells. (**A**) Cells were treated with ionomycin and $Ca^{2+}$ influx was detected. Peak calcium levels are quantified in bar graphs. (**B-C**) Signature protein expression for Th1 (IFNγ), Th17 (IL-17) and iTreg (Foxp3) measured by flow cytometry showed distinct dependencies on SOCE with Th17 cells having the strongest dependency, followed by Th1 cells, while differentiation of iTreg cells is sufficient even with low amounts of calcium influx, n=5-8 per condition, student's t test, *p <0.05, **p<0.01, ***p<0.001

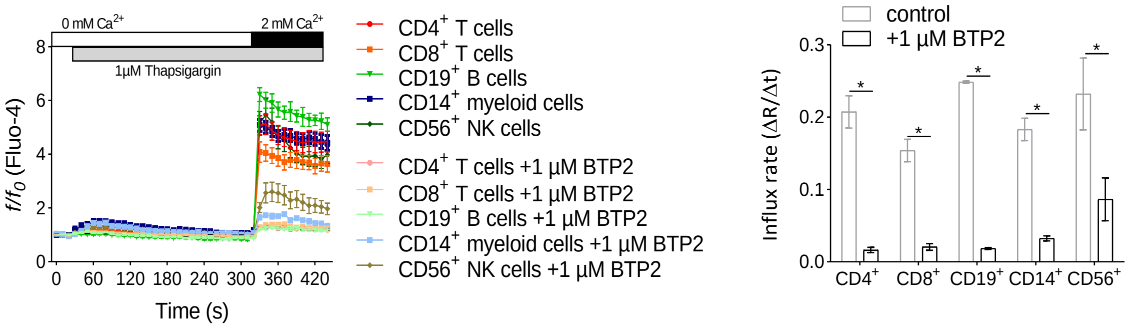

**Supplementary Figure 2**. $Ca^{2+}$ influx analyses were performed by flow cytometry in order to assess $Ca^{2+}$ influx in PBMCs isolated from healthy donors (n = 3), with or without 1 µM BTP2 treatment. *Discovery determined using the two-stage linear step-up procedure of Benjamini, Krieger and Yekutieli, with Q = 1%. Error bars represent the standard error mean (SEM).

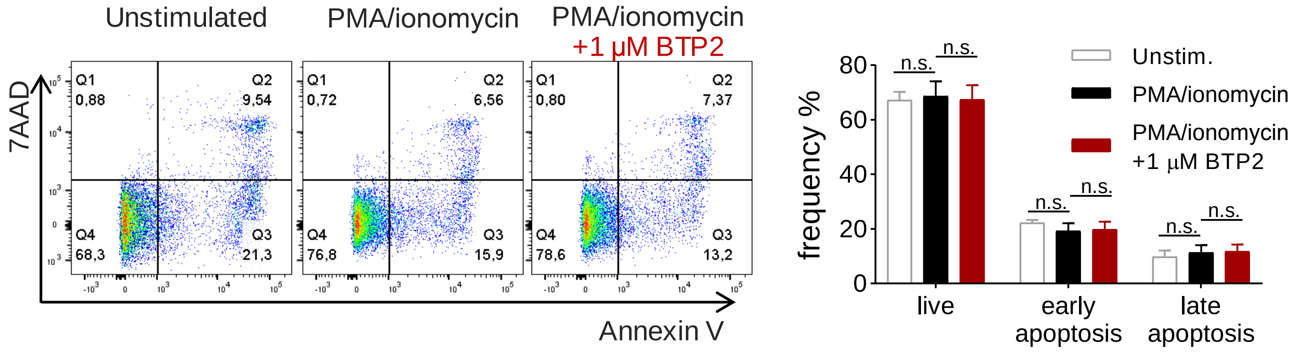

**Supplementary Figure 3**. LPMCs were isolated from 3 IBD patients and cellular viability in the presence or absence of PMA/Ionomycin (4 h in vitro) or additional BTP2 was assessed by quantifying frequencies of early apoptotic (Annexin V$^+$ 7AAD$^-$) and late apoptotic cells (Annexin V$^+$ 7AAD$^+$). Statistic was calculated by paired Wilcoxon matched-pairs signed rank test, *p <0.05. Error bars represent thestandard error mean (SEM).

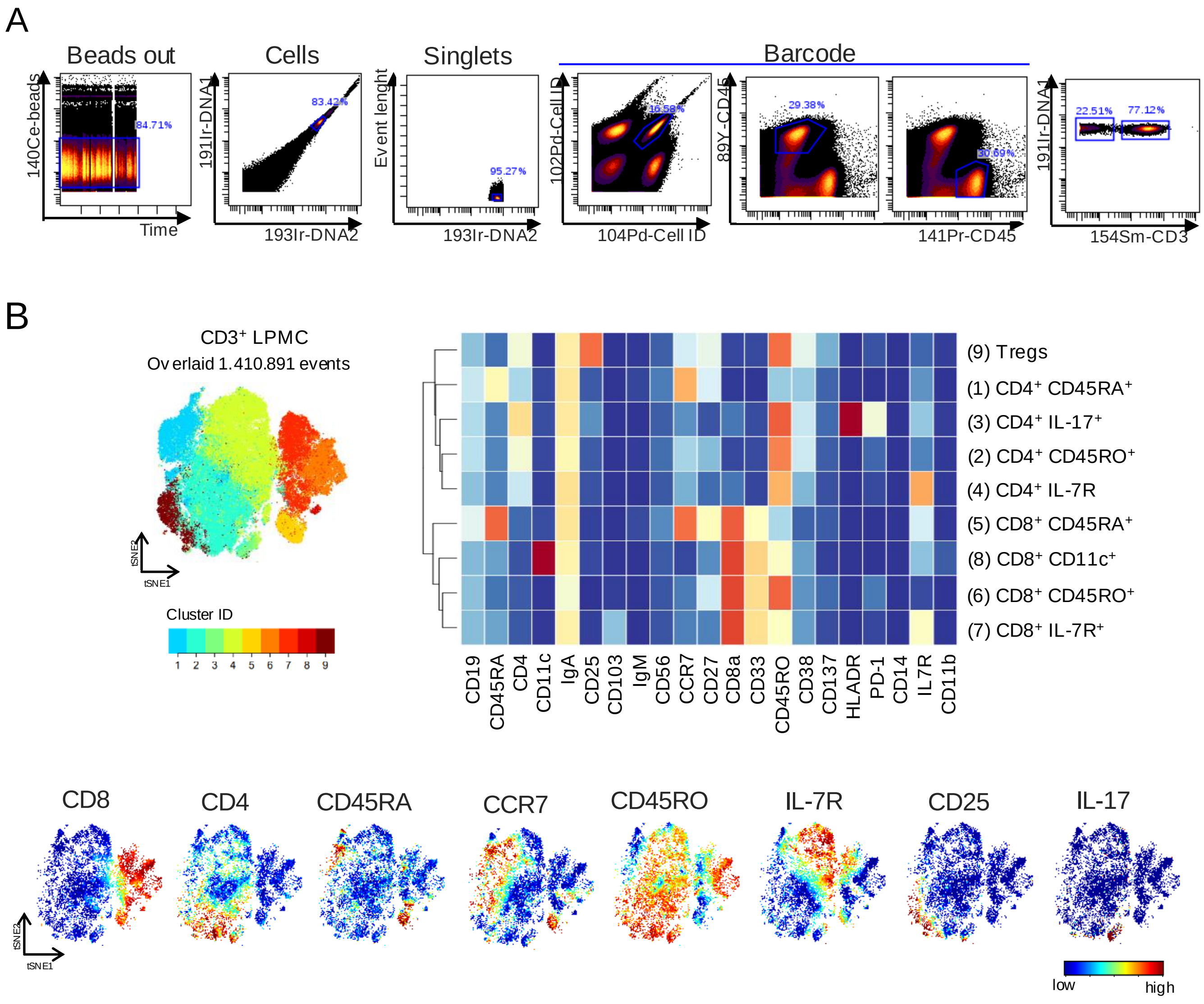

**Supplementary Figure 4**. (**A**) Gating strategy preceding analyses using the t-distributed stochastic linear embedding (t-SNE) algorithm and FlowSOM/ConsensusClusterPlus self-organizing map. Cells were cleaned from calibration beads and doublets and de-barcoded according to the Cell-ID 20-plex Pd Barcoding Kit and 89Y-CD45 staining. (**B**) t-SNE plot of merged FCS files from samples treated with PMA/ionomycin±1μM BTP2 (CD: n =5). Colors on the t-SNE plot indicate 9 defined clusters among $CD45^{+}CD3^{+}$ LPMCs. Heatmap clusters show the expression levels of the 21 markers used for cluster analysis. viSNE plots display representative LPMCs isolated from one CD patient colored by marker expression levels (blue: low, red: high).

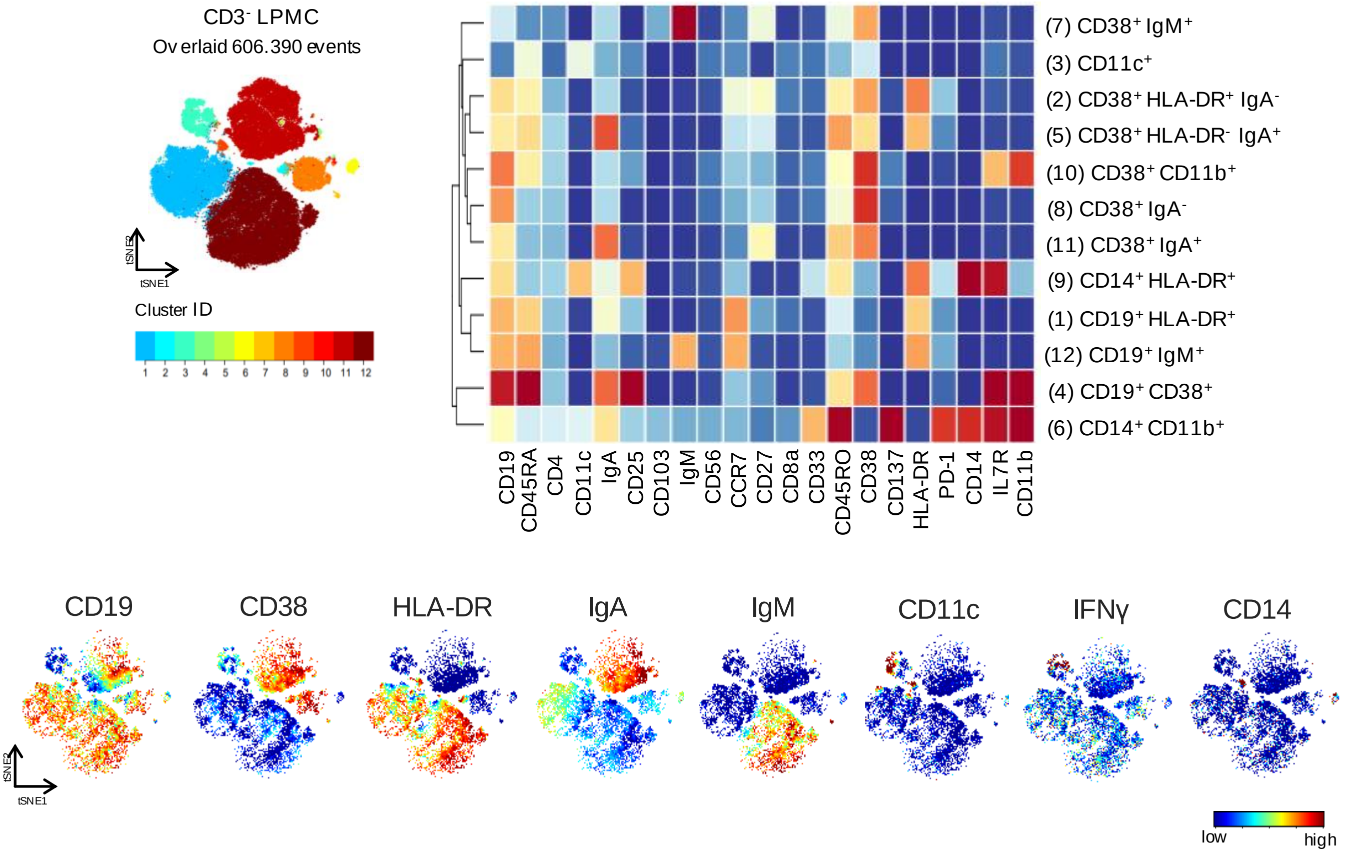

**Supplementary Figure 5**. t-SNE plot of merged FCS files from samples treated with PMA/ ionomycin±1µM BTP2 (CD: n =5). Colors on the t-SNE plot indicate 12 defined clusters among CD45$^{+}$CD3$^{-}$ LPMCs. Heatmap clusters show the expression levels of the 21 markers used for cluster analysis. viSNE plots represent LPMCs isolated from one CD patient colored by marker expression levels (blue: low, red: high).

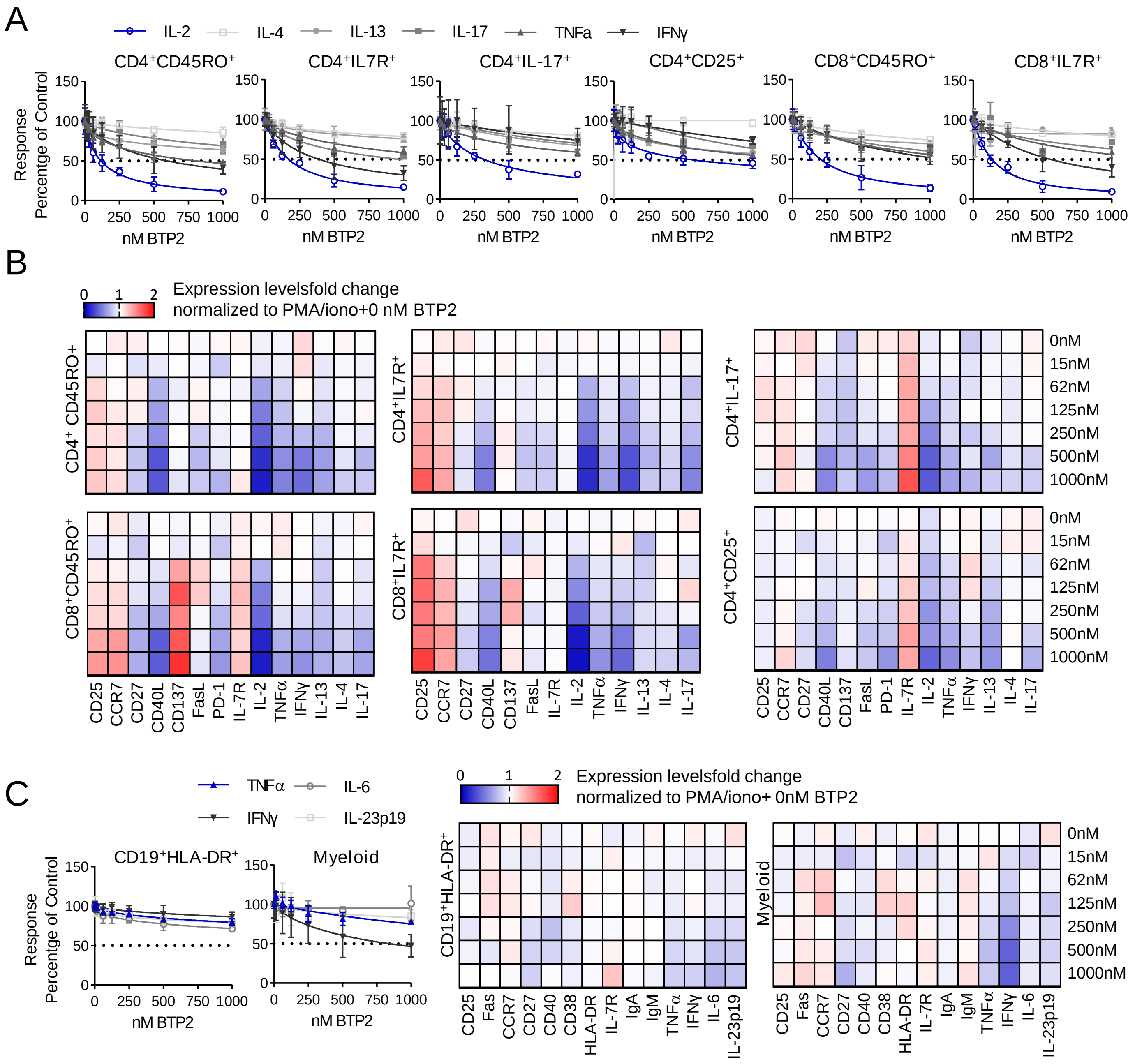

**Supplementary Figure 6.** Dose-dependent suppression of SOCE inhibits the pro-inflammatory cytokine production by lamina propria T cells of UC patients. (**A**) Dose response curves reflecting the normalized production of cytokines in response to treatment with increasing dosages (15-1000nM) of BTP2 in $CD45^{+}CD3^{+}$ T cells after 4 h of ex vivo stimulation with PMA/ionomycin (UC: n=3). The dose response was normalized to control samples treated with PMA/ionomycin. (**B**) Heatmaps representing the median fold change of cytokines and functional markers expression in $CD45^{+}CD3^{+}$ LPMCs (UC: n=3) activated 4 h ex vivo with PMA/ionomycin±15-1000nM BTP2, normalized to samples treated with PMA/Ionomycin. (**C**) Dose response curve graphs reflecting the normalized production of cytokines in response to treatment with increasing dosages (15-1000nM) of BTP2 in $CD45^{+}CD3^{-}$ cells after 4 h of ex vivo stimulation with PMA/ionomycin (UC: n=3). The dose response was normalized to control samples treated with PMA/ionomycin. Heatmaps representing the median fold change of cytokines and functional markers expression in $CD45^{+}CD3^{-}$ LPMCs (UC: n=3) after 4 h of ex vivo stimulatonwith PMA/ionomycin±15-1000nM BTP2, normalized to samples treated with PMA/ionomycin.

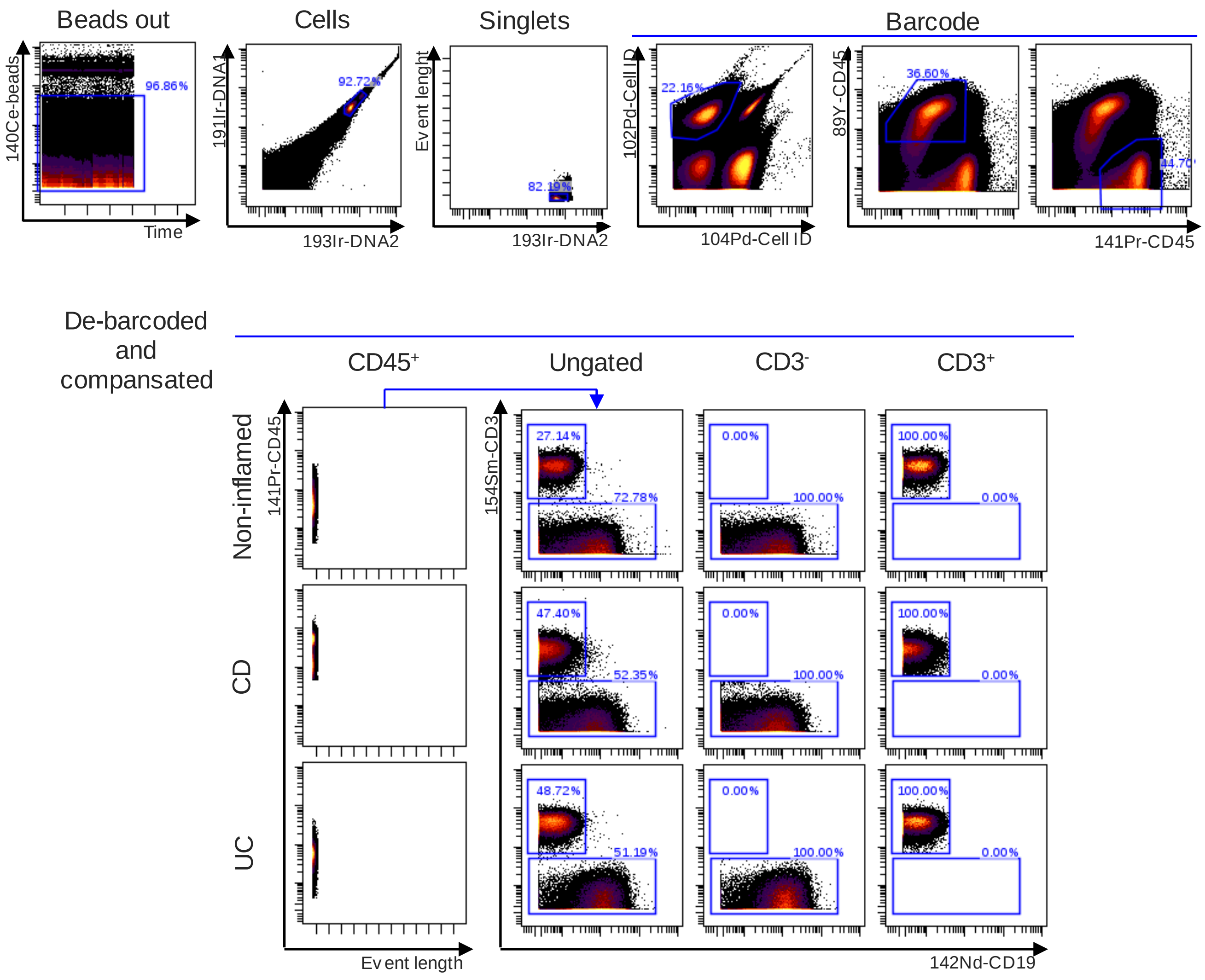

**Supplementary Figure 7.** Gating strategy preceding analyses using the t-distributed stochastic linear embedding (t-SNE) algorithm and FlowSOM/ConsensusClusterPlus self-organizing map. After exclusion of beads and doublets, de-barcoding and compensation, cells were gated on CD45$^+$ or CD45$^+$CD3$^+$ and CD45$^+$CD3$^-$ for further deep immune-characterization.

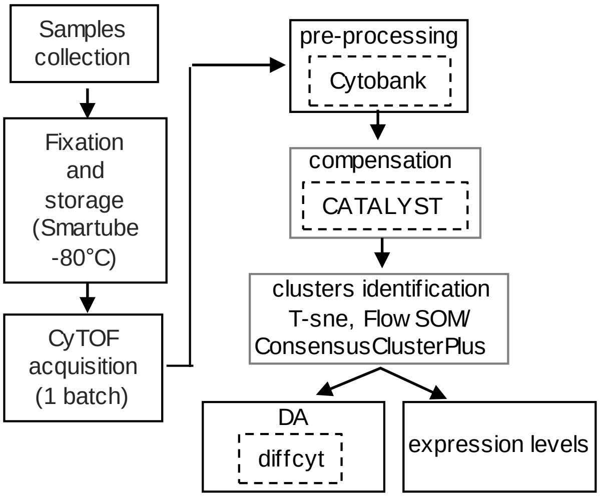

**Supplementary Figure 8.** Experimental design- and analysis workflow applied to CyTOF data.

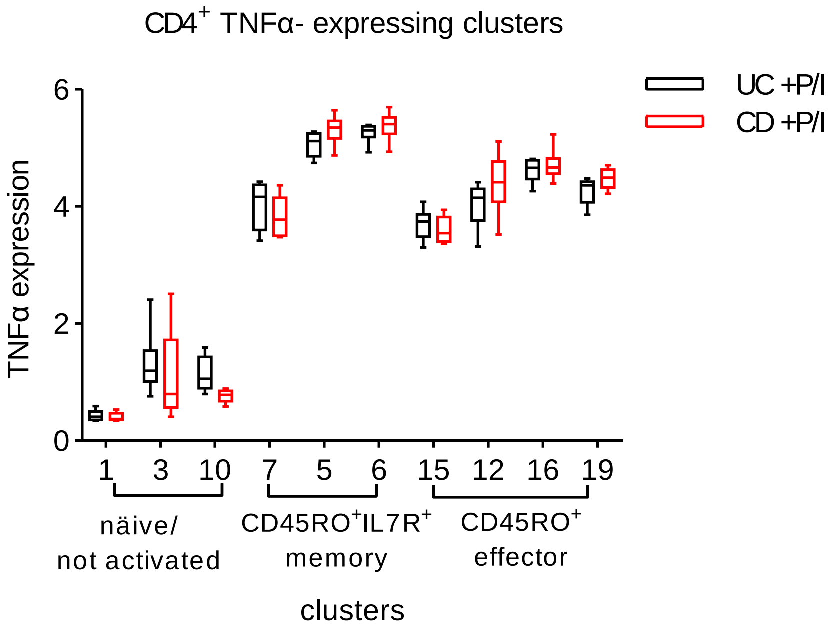

**Supplementary Figure 9.** TNFα expression among $CD4^+$ näive/not activated (clusters 1,3 and 10), memory (clusters 7,5 and 6) or effector T cells (clusters 15,12,16 and 19) identified by the FlowSOM/ConsensusClusterPlus analysis performed on $CD3^+CD45^+$ LPMCs isolated from UC and CD patients and activated for 4 h in vitro with PMA/ionomycin (**Figure 4**). Boxplots showing mean expression levels (arbitrary unit) of TNFα in each cell cluster. Boxes extend from the $25^{th}$ to $75^{th}$ percentiles. Whisker plots show the min (smallest) and max (largest) values. The line in the box denotes the median.

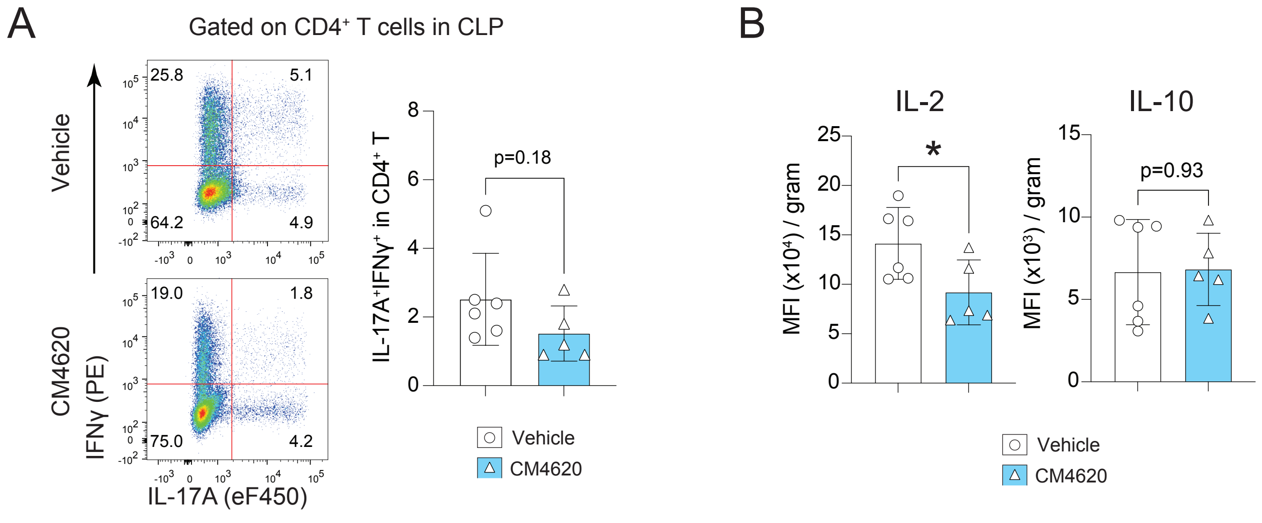

**Supplementary Figure 10.** (**A**) Flow cytometric analysis of IFN-γ and IL-17 producing CD4+ T cells obtained from the colon lamina propria of colitic mice after treatment with vehicle or the SOCE inhibitor CM4620. (**B**) Cytometric bead analyses (CBA) of IL-2 and IL-10 in the supernatant of colon lamina propria derived lymphocytes stimulated with 0.5ug/mL anti-CD3 for 24 hours. Each dot represents one mouse. Statistical analysis by unpaired student's t-test: ***p<0.001 **p<0.01 *p<0.05.

| | |
|---|---|
| ***Total Patients*** | 24 |
| ***Sex*** | |
| Males | 9 |
| Male % | 37.5% |
| Females | 15 |
| Female % | 62.5% |
| ***Age*** | |
| Mean | 45.8 |
| Median | 40.5 |
| Range min | 19 |
| Range max | 91 |
| ***Disease/control*** | |
| Crohn's disease | 11 |
| | 45.8% |
| Ulcerative colitis | 9 |
| | 37.5% |
| Control | 4 |
| | 16.7% |
| ***On ant-- inflammatory*** | 11 |
| ***medication at*** | 45.8% |
| ***inclusion*** | |
| ***Mean leucocyte count per nl at inclusion*** | 7.45 |
| Range min | 4.28 |
| Range max | 16.26 |

**Supplementary Table 1**: Patients‘ characteristics at inclusion.

| Metal | target | Isotype | Clone | Company | dilu3*. |
|---|---|---|---|---|---|
| 89Y | CD45 | IgG1 | HI30 | Fluidigm | 1:100 |
| 141Pr | CD45 | IgG1 | HI30 | Fluidigm | 1:100 |
| 142Nd | CD19 | IgG1 | HIB19 | Fluidigm | 1:100 |
| 143Nd | CD45RA | Mouse IgG2b | HI100 | Fluidigm | 1:100 |
| 144Nd | IL-4 | Rat IgG1 | MP4-25D2 | Fluidigm | 1:100 |
| 145Nd | CD4 | Mouse IgG1 | RPA-T4 | Fluidigm | 1:50 |
| 146Nd | TNF$\alpha$ | IgG1 | Mab11 | Fluidigm | 1:100 |
| 147Sm | CD11c | Mouse IgG1 | Bu15 | Fluidigm | 1:200 |
| 148Nd | IgA | Goat | Polyclonal | Fluidigm | 1:100 |
| 149Sm | CD25 | Mouse IgG1 | 2A3 | Fluidigm | 1:100 |
| 150 Nd | CD86 | IgG2b | IT2.2 | Fluidigm | 1:100 |
| 151Eu | CD103 | Mouse IgG1 | Ber-ACT8 | Fluidigm | 1:100 |
| 152Sm | Fas | Mouse IgG1 | DX2 | Fluidigm | 1:200 |
| 153Eu | IgM | Mouse IgG1 | MHM- 88 | Biolegend | 1:200 |
| 154Sm | CD3 | Mouse IgG1 | UCTH1 | Fluidigm | 1:100 |
| 155Gd | CD56 | IgG1 | B159 | Fluidigm | 1:100 |
| 156Gd | IL- 6 | IgG1 | MQ2-13AS | Fluidigm | 1:100 |
| 158Gd | IFN$\gamma$ | Mouse IgG1 | B27 | Fluidigm | 1:400 |
| 159Tb | CCR7 | IgG2a | G043H7 | Fluidigm | 1:200 |
| 160Gd | CD27 | Mouse IgG1, k | 2EA | Biolegend | 1:200 |
| 161Dy | IL-23p19 | IgG2b | 23dcdp | Fluidigm | 1:100 |
| 162Dy | CD8 | Mouse IgG1 | RPA-T8 | Fluidigm | 1:100 |
| 163Dy | CD33 | Mouse IgG1 | WM53 | Fluidigm | 1:100 |
| 164Dy | CD45RO | Mouse IgG2a | UCHL1 | Fluidigm | 1:100 |
| 165Ho | CD40 | Mouse IgG1 | 5C3 | Fluidigm | 1:100 |
| 166Er | IL- 2 | Rat IgG2a | MQ117H12 | Fluidigm | 1:100 |
| 167Er | CD38 | Mouse IgG1 | HIT2 | Fluidigm | 1:200 |
| 168Er | CD40L | Mouse IgG1 | 24-31 | Fluidigm | 1:100 |
| 169Tm | IL-13 | Rat IgG1 | JES105A2 | Fluidigm | 1:100 |
| 170Er | CD137 | Mouse IgG1, k | 4B4-1 | Biolegend | 1:100 |
| 171Yb | FasL | Mouse IgG1, k | NOK-1 | Biolegend | 1:100 |
| 172Yb | IL-17 | Mouse IgG1 | BL168 | Fluidigm | 1:100 |
| 173Yb | HLA-DR | Mouse IgG2a | L243 | Fluidigm | 1:200 |
| 174Yb | PD-1 | Mouse IgG1 | EH12.2H7 | Fluidigm | 1:200 |
| 175Lu | CD14 | Mouse IgG2a | M5E2 | Fluidigm | 1:50 |
| 176Yb | IL-7R | Mouse IgG1 | A019D5 | Fluidigm | 1:50 |
| 209Bi | CD11b | IgG1 | ICRF44 | Fluidigm | 1:100 |

**Supplementary Table 2.** Antibodies used for Mass Cytometry staining**.**

| Metal | target |
|---|---|
| 142Nd | CD19 |
| 143Nd | CD45RA |
| 145Nd | CD4 |
| 147Sm | CD11c |
| 148Nd | IgA |
| 149Sm | CD25 |
| 151Eu | CD103 |
| 153Eu | IgM |
| 155Gd | CD56 |
| 159Tb | CCR7 |
| 160Gd | CD27 |
| 162Dy | CD8 |
| 163Dy | CD33 |
| 164Dy | CD45RO |
| 167Er | CD38 |
| 170Er | CD137 |
| 173Yb | HLA-DR |
| 175Lu | CD14 |
| 176Yb | IL- 7R |
| 209Bi | CD11b |

**Supplementary Table 3.** Markers used for viSNE clustering of $CD45^{+}CD3^{+}$ and $CD45^{+}CD3^{-}$ LPMCs described in **Figure 2D-G.**

| Metal | target |
|---|---|
| 142Nd | CD19 |
| 143Nd | CD45RA |
| 145Nd | CD4 |
| 147Sm | CD11c |
| 148Nd | IgA |
| 149Sm | CD25 |
| 151Eu | CD103 |
| 153Eu | IgM |
| 154Sm | CD3 |
| 155Gd | CD56 |
| 159Tb | CCR7 |
| 160Gd | CD27 |
| 162Dy | CD8 |
| 163Dy | CD33 |
| 164Dy | CD45RO |
| 167Er | CD38 |
| 170Er | CD137 |
| 173Yb | HLA- DR |
| 175Lu | CD14 |
| 176Yb | IL-7R |
| 209Bi | CD11b |

**Supplementary Table 4.** Markers used for viSNE clustering of $CD45^+$ LPMCs described in **Figure 3.**

| Metal | target |
|---|---|
| 143Nd | CD45RA |
| 144Nd | IL-4 |
| 145Nd | CD4 |
| 146Nd | TNF$\alpha$ |
| 149Sm | CD25 |
| 151Eu | CD103 |
| 152Sm | Fas |
| 156Gd | IL-6 |
| 158Gd | IFNg |
| 159Tb | CCR7 |
| 160Gd | CD27 |
| 161Dy | IL- 23p19 |
| 162Dy | CD8 |
| 164Dy | CD45RO |
| 165Ho | CD40 |
| 166Er | IL- 2 |
| 167Er | CD38 |
| 168Er | CD40L |
| 169Tm | IL-13 |
| 170Er | CD137 |
| 171Yb | FasL |
| 172Yb | IL-17 |
| 174Yb | PD-1 |
| 176Yb | IL-7R |

**Supplementary Table 5.**Markers used for viSNE clustering of CD45$^+$CD3$^+$ LPMCs described in **Figure 4.**

| ***Metal*** | ***target*** |
|---|---|
| *142Nd* | *CD19* |
| *143Nd* | *CD45RA* |
| *144Nd* | *IL-4* |
| *145Nd* | *CD4* |
| *146Nd* | *TNF$\alpha$* |
| *147Sm* | *CD11c* |
| *148Nd* | *IgA* |
| *149Sm* | *CD25* |
| *150 Nd* | *CD86* |
| *151Eu* | *CD103* |
| *152Sm* | *Fas* |
| *153Eu* | *IgM* |
| *155Gd* | *CD56* |
| *156Gd* | *IL-6* |
| *158Gd* | *IFN$\gamma$* |
| *159Tb* | *CCR7* |
| *160Gd* | *CD27* |
| *161Dy* | *IL-23p19* |
| *162Dy* | *CD8* |
| *163Dy* | *CD33* |
| *164Dy* | *CD45RO* |
| *165Ho* | *CD40* |
| *166Er* | *IL-2* |
| *167Er* | *CD38* |
| *168Er* | *CD40L* |
| *169Tm* | *IL-13* |
| *170Er* | *CD137* |
| *171Yb* | *FasL* |
| *172Yb* | *IL-17* |
| *173Yb* | *HLA-DR* |
| *174Yb* | *PD-1* |
| *175Lu* | *CD14* |
| *176Yb* | *IL-7R* |
| *209Bi* | *CD11b* |

**Supplementary Table 6**. Markers used for viSNE clustering of $CD45^{+}CD3^{-}$ LPMCs described in **Figure 5.**

| Antigen | Manufacturer | Clone# | Conjugation |
|---|---|---|---|
| CD11b | eBioscience | M1/70 | PE-Cy7 |
| CD4 | Biolegend | GK1.5 | APC-Cy7 |
| CD44 | eBioscience | IM7 | FITC |
| CD62L | Biolegend | Mel-14 | PerCP-Cy5.5 |
| IFN-γ | Biolegend | XMG1.2 | PE |
| IL-17A | eBioscience | eBio17B7 | eFluor 450 |
| IL-2 | eBioscience | JES6-5H4 | FITC |
| TNF-α | eBioscience | MP6-XT22 | APC |
| Ly6G | Biolegend | 1A8 | Alexa-Fluor647 |
| CD25 | Biolegend | PC61 | PE |
| Foxp3 | eBioscience | FJK-16s | PE |
| RORgt | eBioscience | B2D | APC |

**Supplementary Table 10.** Antibodies used for Flow Cytometry staining in T cell transfer models of colitis**.**
